# Extrachromosomal DNA associates with poor survival across a broad spectrum of childhood solid tumors

**DOI:** 10.1101/2025.07.22.24308163

**Authors:** Owen S. Chapman, Sunita Sridhar, Eugene Yui-Ching Chow, Rishaan Kenkre, Jonathan Kirkland, Aditi Dutta, Shanqing Wang, Riki Goto, Wenshu Zhang, Miguel Brown, Jens Luebeck, Hui Hui, Jessica Wang, Yan Yuen Lo, Elias Rodriguez-Fos, Shanzheng Wang, Konstantin Okonechnikov, David R. Ghasemi, Kristian W. Pajtler, Johannes Gojo, Anton G. Henssen, Marcel Kool, Vineet Bafna, Daisuke Kawauchi, Megan Paul, Kevin Yip, Jill P. Mesirov, Lukas Chavez

## Abstract

Circular extrachromosomal DNA (ecDNA) is a powerful driver of oncogene amplification and tumor evolution, yet its prevalence, composition, and clinical significance across pediatric cancers remains incompletely understood. Leveraging two major cloud-based genomic data repositories, we analyzed whole genome sequencing data from 3,630 tumor biosamples representing 2,967 children across 39 solid tumor types. ecDNA was identified in 9% of cases and was enriched in high-grade and clinically aggressive malignancies including ETMR, pediatric high-grade glioma, medulloblastoma, neuroblastoma, osteosarcoma and rhabdomyosarcoma. We catalogued 392 ecDNA sequences, revealing recurrent amplification of known oncogenes, diverse gene fusions, and oncogenic loci where recurrent ecDNA involvement underscores their emerging importance in pediatric tumors. Oncogenes amplified on ecDNA reached significantly higher copy number than chromosomal amplifications, and ecDNA was associated with significantly poorer 5-year survival independent of tumor type, age and sex. Longitudinal analyses demonstrated that ecDNA frequently arises, is lost, or undergoes structural remodeling during progression and recurrence, including acquisition or loss of oncogenes on the same circular element. Together, these findings define the landscape, clinical relevance, and evolutionary behavior of ecDNA across childhood cancers, highlight candidate drivers, and identify patient populations that may benefit from emerging ecDNA-targeted therapeutic strategies. An interactive resource is available at https://ccdi-ecdna.org/.

## INTRODUCTION

Extrachromosomal DNA (ecDNA, also known as double minutes), are megabase-scale circular focal amplifications of genomic DNA which arise in many human cancers. ecDNA sequences generally contain an oncogene or other functional element conferring a selective fitness advantage, and lack a centromere, leading to unequal segregation during cell division and intra-tumoral heterogeneity. The potent combination of copy number heterogeneity and positive selection drives high-copy oncogenic amplification, rapid tumor evolution, and therapeutic resistance^1–3^.

Paired-end whole genome sequencing has emerged as a cost-effective and efficient method to identify oncogenes and other sequences captured on ecDNA. While it was largely overlooked in early pediatric pancancer genomics studies^4,5^, recent studies have described the prevalence of ecDNA in various adult and some childhood cancer types^6–10^. Recognizing that pediatric cancers have 14-fold lower somatic mutation frequency than adult cancers as well as unique drivers and amplifications^4,5^, we have now comprehensively examined the frequency of ecDNA and its association with survival across a broad spectrum of primary and recurrent pediatric cancers.

## RESULTS

### Cloud genomic data repositories facilitate analysis of rare pediatric cancers

To identify ecDNA in pediatric cancers, we analyzed whole genome sequencing (WGS) data available in two large pediatric cancer cloud genomics platforms: the Pediatric Brain Tumor Atlas (PBTA)^11^ and St. Jude Cloud (SJC)^12^ (**Figure 1**). In total, this retrospective cohort comprised 3,630 solid tumor biosamples from 2,967 patients with mean WGS coverage of 69x (±15) per sample. Clinical metadata, including histological diagnoses and patient survival outcomes, were collated from both platforms (**Table 1; Supplementary Table 1**). For brain tumors included in the PBTA, histological diagnoses were integrated with molecular tumor type classifications derived from methylation analyses^13^ of the same samples. We applied the AmpliconArchitect (AA)^14^ and AmpliconClassifier (AC)^15^ algorithms to identify ecDNA amplifications. Briefly, this pipeline performs joint analysis of genomic copy number and structural variation in WGS data to construct a localized genome graph capturing the amplified regions and identify ecDNA as genome cycles satisfying other heuristics including length and copy number (**Methods**). In total, 392 ecDNA sequences were reported in 321 tumor biosamples of 269 patients (9% of patients, **Supplementary Tables 1-3**). No association was observed between presence of ecDNA and age at diagnosis (𝑅^2^ = 0.005, 𝑝 = 0.4, Student’s *t*-test; **Supplementary Figure 1**) or sex (𝜒^2^ = 1.3, 𝑝 = 0.3). As a quality control measure, we estimated tumor purity and found no significant association with the identification of ecDNA (likelihood ratio test, *p* = 0.15, *n* = 2133). Given that previous studies using similar methods did not find ecDNA in adult hematologic malignancies^6,8^, we additionally sampled 10% of available pediatric hematologic malignancies (*n* = 63, **Supplementary Table 4**). No ecDNA was detected in those samples, and we therefore limited the scope of our investigation to solid tumors.

**Figure 1:**
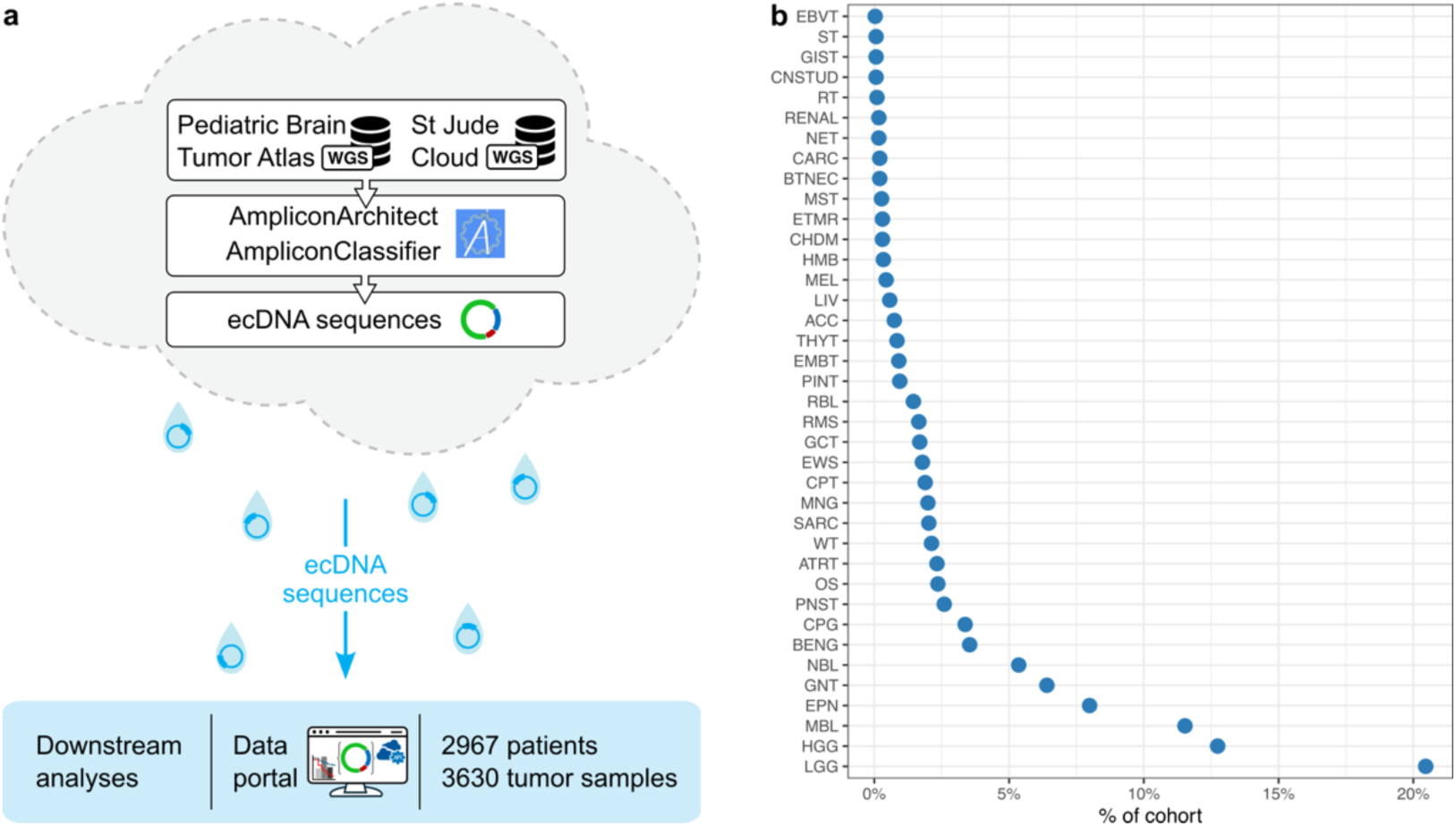
Analysis of ecDNA in pediatric cancers using cloud computing. (**a**) Whole genome sequencing (WGS) data of solid tumors was analyzed in two cancer genome cloud repositories. ecDNA and other amplifications were identified and annotated with clinical information for downstream analysis. (**b**) Tumor biosample distribution by tumor type. Abbreviations are expanded in **Supplementary Table 12**.

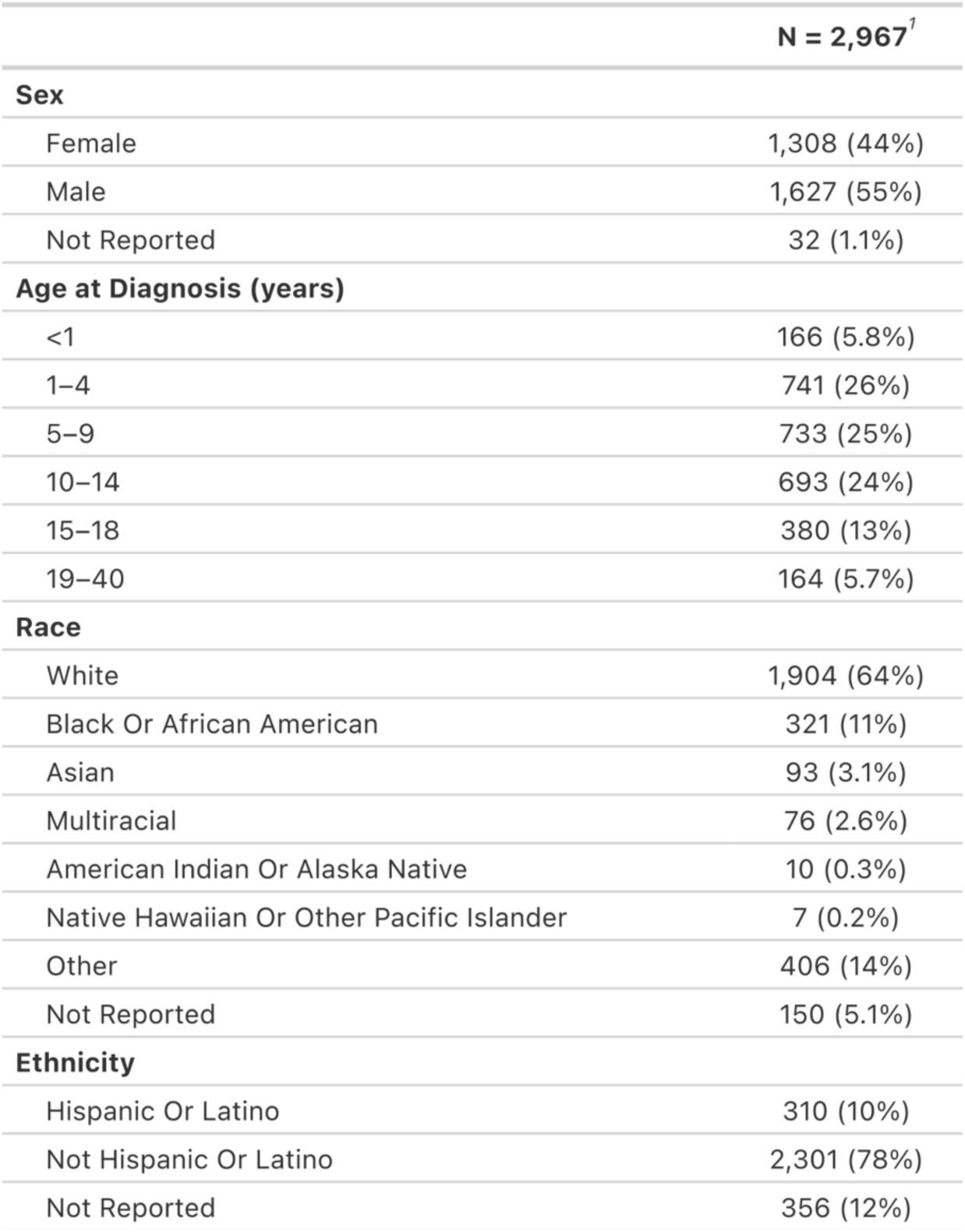

### ecDNA amplifies known and putative oncogenic sequences in pediatric solid tumors

To identify oncogenic drivers amplified on ecDNA in pediatric solid tumors, we defined recurrent genomic intervals as those amplified on ecDNA in at least three independent cases, corresponding to a 99.9% confidence level against chance occurrence (**Supplementary Note 1**; **Supplementary Figure 2**). 109 distinct genomic intervals were recurrently amplified on ecDNA across all pediatric cancers in this cohort (**Figure 2a**). To identify putative driver genes, we annotated these genomic intervals with oncogenes listed in the COSMIC^16^ and ONGene^17^ databases (**Supplementary Table 5**). Genomic intervals most frequently amplified on ecDNA contained oncogenes *MYCN* (66 ecDNA amplifications observed in eight tumor types) or *CDK4* (22 ecDNA amplifications in six tumor types; **Figure 2b**). Five contiguous intervals recurrently amplified on ecDNA contained clusters of three or more oncogenes: chr4q12 (*PDGFRA*, *KIT*, and *KDR*), chr8q24 (*TRIB1*, *MYC*, *PVT1*), chr11q22 (*YAP1*, *BIRC3*, *BIRC2*, *MMP12*), chr12q13-14 (*GLI1*, *DDIT3*, *AGAP2*, *CDK4*), and chr12q15 (*IFNG*, *MDM2, YEATS4*).

**Figure 2:**
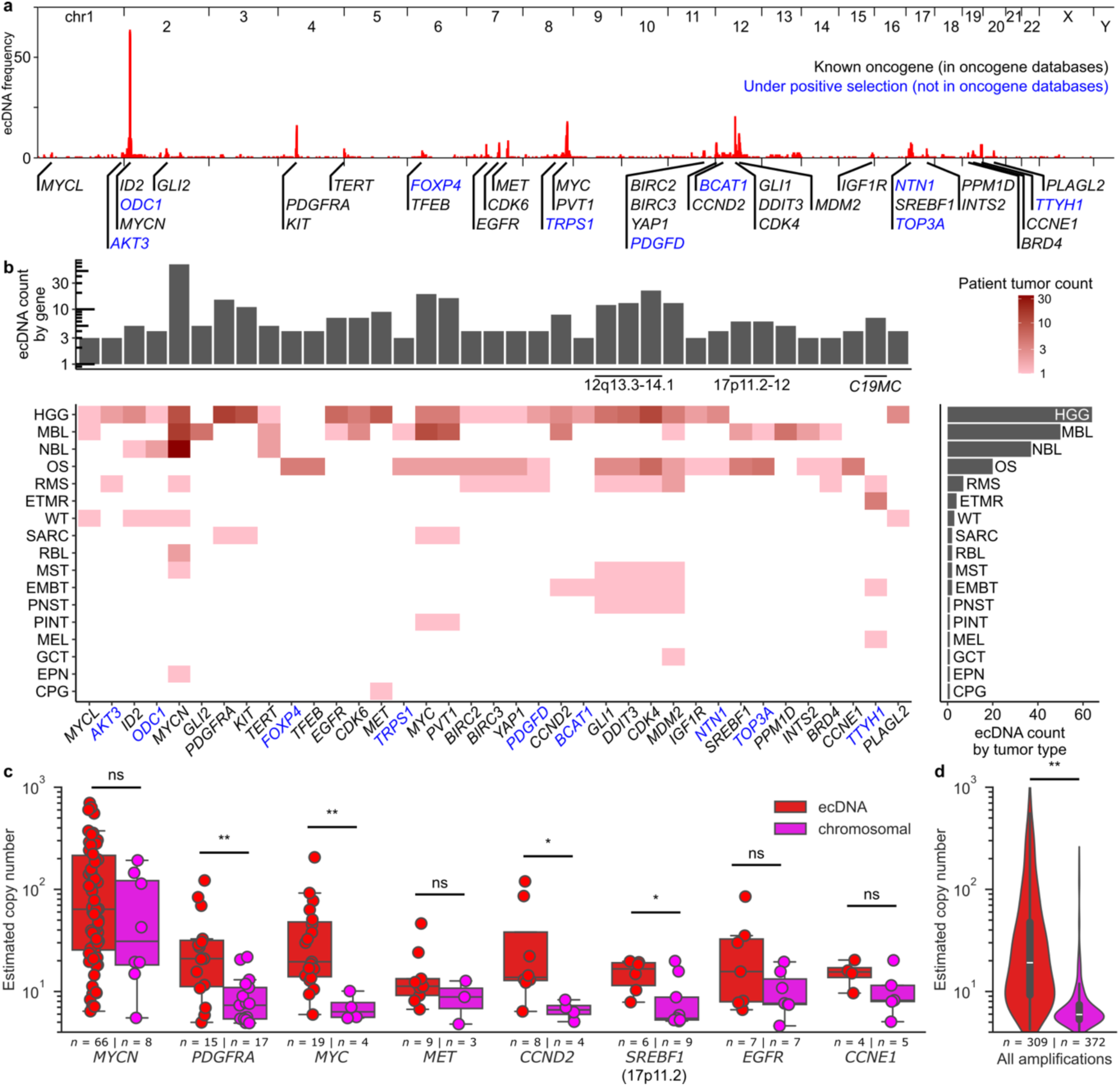
Genomic distribution and copy number of recurrent ecDNA amplifications in pediatric tumors. (**a**) Histogram of ecDNA amplifications in pediatric tumors across the reference human genome. 106 contiguous genomic loci were recurrently (𝑛 ≥ 3) amplified containing 49 different oncogenes. Selected genes are shown: black, oncogenes listed in the COSMIC or ONGene databases; blue, other recurrently amplified genes. (**b**) Amplifications in (**a**) by tumor type. Heatmap indicates the number of cases with ecDNA amplification of a given gene in that tumor type. Right barplot indicates the number of ecDNA sequences amplifying any of the indicated genes for each cancer type. Top barplot indicates the number of ecDNA sequences amplifying the indicated gene across all tumor types. (**c**) Estimated copy number of extrachromosomal and chromosomal amplifications of selected oncogenes. ** 𝑞 < 0.005; * 𝑞 < 0.05; ns, 𝑞 ≥ 0.05; one-sided Mann-Whitney U test with Benjamini-Hochberg FDR correction. (**d**) Estimated copy number of amplicons containing at least 1 gene. Legend and statistical tests are the same as in (c). HGG, high-grade glioma; MBL, medulloblastoma; NBL, neuroblastoma; OS, osteosarcoma; RMS, rhabdomyosarcoma; ETMR, embryonal tumor with multilayered rosettes; WT, Wilms tumor; EMBT, miscellaneous embryonal brain tumors; GCT, germ cell tumors; MST, metastatic secondary tumors; RBL, retinoblastoma; CPG, craniopharyngioma; EPN, ependymoma; MEL, melanoma; PINT, pineal tumors; PNST, peripheral nerve sheath tumors; SARC, other miscellaneous sarcomas.

Accumulating evidence suggests that ecDNA enables greater oncogene copy number amplification than chromosomal amplification^8^. To determine whether amplification by ecDNA is of greater magnitude than other amplifications, we compared estimated copy number for oncogenes amplified in cases on ecDNA and chromosomal amplifications (**Figure 2c**). Median ecDNA copy number was greater for *PDGFRA*, *MYC*, *CCND2*, and chr17p11.2 compared to chromosomal amplification of the same genes in other tumors (one-sided Mann-Whitney *U* with Benjamini-Hochberg FDR correction, *q* < 0.05). Median ecDNA copy number was also greater across all amplifications classified as ecDNA compared to chromosomal (*U* = 1e5; *p* = 2e-61; **Figure 2d**), suggesting that oncogenic amplification occurs at greater magnitude in ecDNA than in chromosomal amplifications in pediatric cancers.

82 genomic intervals recurrently amplified on ecDNA contained no known oncogene. These were relatively short (mean 181kbp, standard deviation 301kbp); however, 38 of these contained uninterrupted gene sequences, nominating 81 protein-coding genes and 235 long noncoding RNAs (lncRNA) as candidates under positive selection (**Supplementary Table 6**). These other recurrently amplified genes exhibited median ecDNA copy numbers comparable to known oncogenes (two-sided Mann-Whitney test; *U* = 138; *p* = 0.23; **Supplementary Figure 3**). Several recurrently amplified genomic intervals were exclusively co-amplified on ecDNA with known oncogenes on recombinant DNA sequences, suggesting that possible positive selection may be context-dependent^18^. To nominate *bona fide* oncogenic candidates, we therefore further asked which genomic intervals were recurrently amplified on ecDNA yet lacked any co-amplified known oncogenes. As a result, we identified five genomic intervals that fulfilled these criteria, each containing genes with prior evidence supporting tumorigenic roles. For example, a peritelomeric recurrently amplified locus at chromosome 1q amplified the full gene sequences of *AKT3* and *ZBTB18* in two high-grade gliomas and a rhabdomyosarcoma (**Supplementary Figure 4**). In mouse models, amplification and overexpression of the paralogous gene *Akt3* promotes malignant glioma progression^19,20^, supporting a functional role for this gene locus in some human pediatric tumors. The other examples contain the DNA replication initiation cofactor *TRPS1*^21^, neurodevelopmental factor *NTN1*^22,23^, the *C19MC* microRNA cluster frequently amplified in embryonal tumors with multilayered rosettes (ETMR)^24^, and the recombination hotspot at chr17p11.2-12 in osteosarcomas^25^ and medulloblastomas^26^.

### ecDNA in aggressive childhood central nervous system tumors

We observed ecDNA most frequently in embryonal tumors with multilayered rosettes (4/9, 44%), pediatric high-grade gliomas (78/391, 20%), and medulloblastomas (55/347, 16%); and rarely in pineal tumors (1/28, 4%), choroid plexus tumors (1/56; 2%), craniopharyngiomas (1/101, 1%), ependymomas (3/239, 1%), glioneuronal tumors (1/192, 0.5%), and low-grade gliomas (1/616, 0.2%) (**Figure 3a**).

**Figure 3:**
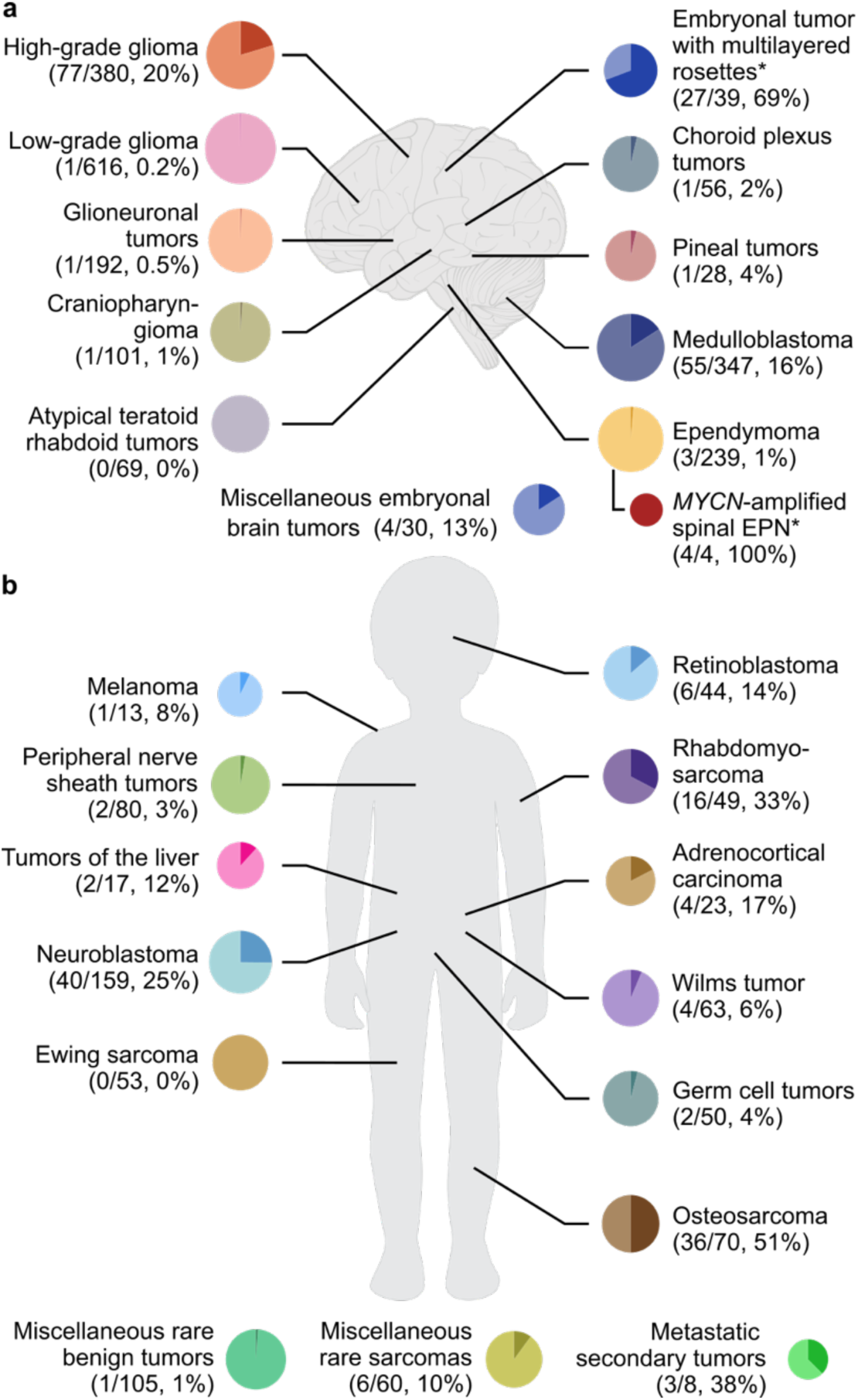
Prevalence of ecDNA across pediatric tumor types. Across 39 pediatric tumor categories, we report ecDNA in 23. Pie chart area is proportional to the log of the sample size for a given tumor. *Includes additional samples from an external validation set analyzed with different methods (see **Methods**).

Embryonal tumors with multilayered rosettes (ETMR) have poor outcomes and most are characterized by amplification of a microRNA cluster on chromosome 19 (*C19MC*) including fusion of the *TTYH1* gene^27^. Of 9 ETMR cases, 4 had *TTYH1*::*C19MC* amplification, all extrachromosomal, suggesting that *C19MC* amplification in ETMR tumors may occur predominantly on ecDNA.

To evaluate this hypothesis, we examined a validation cohort of 30 additional ETMR tumors (**Supplementary Table 7**). Copy number estimation from bulk WGS indicated *C19MC* amplification (CN > 4.5) in 18 cases and moderate gain (2.5 < CN < 4.5) in an additional 8 cases. Using default analysis parameters, 16 of 18 amplifications were classified as ecDNA and 2 as chromosomal amplifications. To improve detection sensitivity, we reanalyzed the 8 cases with moderate *C19MC* gain (**Methods**) and identified an additional 5 ecDNA and 1 chromosomal amplicon, indicating that ecDNA detection sensitivity may be limited for amplicons with low copy burden.

Overall, ecDNA containing *TTYH1*::*C19MC* fusion was detected in 21 of 30 cases in the validation cohort (70%). Chromosomal amplicons containing the fusion were detected in an additional 3 cases (10%). The fraction of *C19MC* amplicons classified as extrachromosomal in the validation set (21/24) did not differ significantly from the discovery set (4/4; *p* = 1, Fisher exact test), substantiating the high prevalence of extrachromosomal *C19MC* amplification in a larger ETMR cohort. Moreover, ecDNA amplifications of *KANSL1* and *YAP1* were found in non-*C19MC*-amplified tumors in the validation set, suggesting possible alternative driver amplifications of non-*C19MC*-amplified ETMR.

The clinical outcomes of pediatric high-grade glioma (pHGG) patients differ across molecular subgroups marked by genetic mutations to the histone H3 or isocitrate dehydrogenase (*IDH*) genes^28^. pHGG with H3 mutations are further stratified into those mutated at lysine 27 (H3K27) and at glycine 34 (H3G34). Among 380 pHGG tumors, 77 had ecDNA (20%), including 29 of 103 with wild-type H3 (28%), 29 of 132 H3K27-mutant (22%), 2 of 16 H3G34-mutant (13%), and 4 of 10 with *IDH* mutation (40%). Known oncogenes^28–30^ recurrently amplified on ecDNA included *PDGFRA* (*n* = 14), *CDK4* (12), *MYCN*, *KIT* (10), *MET* (8), *EGFR* (6), *GLI1* (5), *CDK6, MYC, MDM2* (4), *ID2, CCND2,* and *PLAGL2* (3).

In a previous analysis of 481 medulloblastoma (MBL) tumors, we showed significant associations between ecDNA status and survival across all MBL tumors and in each molecular subgroup^7^. 168 MBL tumors were reanalyzed herein alongside 174 new cases. ecDNA was detected in 16%, modestly lower than that reported previously (18%), which may be attributed to differences in the patient cohorts and analytical methods (**Supplementary Note 2**).

The low frequency of ecDNA detected in choroid plexus tumors, pineal tumors, craniopharyngiomas, ependymomas, CNS glioneuronal tumors, and low-grade gliomas (**Supplementary Note 3**) prompts the question of whether ecDNA amplifications are incidental findings or characteristic to well defined rare molecular subtypes. For pineal tumors and ependymomas with available DNA methylation-based classifications, ecDNA amplifications were indeed limited to the recently-described rare molecular subtypes *MYC-*amplified pineoblastoma^31^ and *MYCN*-amplified spinal ependymoma^32,33^ respectively. To validate these findings, we obtained new WGS data from 3 *MYCN*-amplified spinal ependymomas. Amplicon analysis identifies ecDNA in each, suggesting that amplification of *MYCN* in this molecular subtype may commonly be driven by ecDNA. Overall, although further studies of larger tumor-specific cohorts are needed, these observations suggest that ecDNA amplification may be limited to some ultra-rare and highly aggressive subtypes and are otherwise largely absent in these tumor types.

### ecDNA in pediatric extracranial solid tumors

ecDNA has been previously described in various extracranial pediatric solid tumors, most extensively in neuroblastoma^9,18^, and in case reports and cell lines of retinoblastoma^34,35^, rhabdomyosarcoma^29,36–41^, and osteosarcoma^42^. We identify patient tumors with ecDNA for each of these extracranial tumor types, as well as in adrenocortical carcinomas, Wilms tumors, germ cell tumors, malignant peripheral nerve sheath tumors, and other rare sarcomas (**Figure 3b**).

Consistent with previous analyses^9,10^, we estimate the prevalence of ecDNA in neuroblastoma at 40/159 (25%). Most tumors with ecDNA amplified *MYCN* (35/40, 88%). Of 5 remaining ecDNA(+) tumors not amplifying *MYCN*, two had ecDNA amplifications of *TERT*, and 3 had low-copy ecDNA containing no known oncogenic sequences.

Although *RB1* gene inactivation is the canonical driver of retinoblastoma, other mechanisms including oncogenic amplification are believed to contribute to tumorigenesis in some cases^43^. ecDNA has previously been described in retinoblastoma cell lines and patient tumors and is particularly associated with *MYCN* amplification^35^. In this cohort, six of 44 (14%) retinoblastoma cases had ecDNA, two of which amplified *MYCN*. A further two ecDNA amplifications disrupted the *RB1* gene to form gene fusions, with lipoma *HMGIC* fusion partner gene family member *LHFPL6* (*RB1-LHFPL6)* or with the *TPTE2P2* pseudogene (*RB1-TPTE2P2)*. In the remaining two tumors, no canonical oncogenes were identified, suggesting that these highly amplified ecDNA sequences may contain previously unknown oncogenic drivers for this tumor type.

Osteosarcomas are frequently characterized by high genomic instability with some genomic regions frequently involved in copy number gain events^44,45^. Prognosis is poor with few advances in standard treatment for the past 30 years^46^. ecDNA was detected in 36 of 70 cases (51%). We identified recurrent ecDNA amplifications at chromosomes 6p21 (*FOXP4*^47^, 4 cases), 12q13 (*GLI1*, *CDK4*, 6 cases), 17p11.2-p12^48,49^ (11 cases), and 19q12 (*CCNE1*, 4 cases). 3 tumors had more than one of these recurrently amplified regions on distinct ecDNA sequences.

Double minutes have been described in case reports of rhabdomyosarcoma (RMS) as early as 1971^36,37,39,50,51^. Amplifications of *MYCN*^52^ and fusion oncogenes between *FOXO1* and *PAX3* or *PAX7*^53^ have been previously identified as prognostic indicators. ecDNA was detected in 16 of 49 RMS cases (33%). Amplified oncogenes included *PAX7*-*FOXO1* fusion, *MDM2* (*n* = 2), *MYCN*, *FGFR1*, *NCOA1*-*PAX3* fusion, and *NSD3* (*n* = 1). One tumor harbored ecDNA where none of the amplified genes were annotated in the two queried oncogene databases; however, we suggest the cell cycle regulators *CDK11A* and *CDK11B* as likely oncogenic candidates.

In contrast to a previous pan-cancer study^6^, we did not observe ecDNA in any of 53 pediatric Ewing sarcomas. On further investigation, we noted that tumors previously annotated as Ewing sarcomas have been subsequently reannotated as osteosarcomas (**Supplementary Table 8**), explaining the discrepancy and suggesting that ecDNA does not play a major role in oncogenesis of pediatric Ewing sarcomas.

We also report ecDNA sequences in 4 adrenocortical carcinomas, 4 Wilms tumors, 2 germ cell tumors, 2 malignant peripheral nerve sheath tumors, 2 hepatoblastomas, one pediatric melanoma, and one hepatic focal nodular hyperplasia. These cases are described in **Supplementary Note 4**.

### Tumors with ecDNA, chromosomal, or no focal amplification have distinct distributions of somatic small variants

Somatic variant calling and annotation were performed on paired blood/tumor WGS data from both the SJ and CBTN datasets to assess whether the distribution of somatic mutation (single nucleotide variants (SNVs) and short indels < 50bp) differs in ecDNA-amplified tumors. For this analysis, we defined likely pathogenic (*LP*) somatic variants as somatic mutations of cancer-related genes which altered the protein sequence and were predicted to be functional (**Methods**). Aggregating *LP* mutations at the gene level, we first compared amplified to nonamplified tumors (**Supplementary Figure 5a**). Mutations of *BRAF* and *CTNNB1* were depleted in tumors with amplification, likely reflecting enrichment of *BRAF* and *CTNNB1* mutations in low-grade gliomas, glioneuronal tumors, and *WNT* subgroup medulloblastomas in which focal amplification is rare. Amplified tumors were enriched for mutation of *TP53*, *H3-3A*, *KNL1*, *PDGFRA*, *PABPC1*, *AURKA*, *STAG2*, and *KNSTRN* (*n* = 7071 *LP* somatic variants across 1798 solid tumors; *χ*^2^ tests with FDR correction, *q* < 0.10; **Supplementary Table 9**). Notably, *KNL1*, *AURKA, STAG2* and *KNSTRN* are required for proper chromosomal pairing and segregation during mitosis, suggesting a plausible link between defects in this process and the presence of focal amplification in pediatric solid tumors. Due to the large hypothesis space, this analysis was underpowered to examine differences between extrachromosomally and chromosomally amplified tumors, but we note that alterations to *GNAS* were nominally depleted in tumors with ecDNA vs. amplified tumors without ecDNA (**Supplementary Figure 5b**). In an analysis of germline variation using similar methods across a subset of CNS tumors, pathogenic germline variants of *TP53* (Li-Fraumeni Syndrome, LFS) were associated with focal amplification overall but did not show specific enrichment for ecDNA (**Supplementary Note 5**).

We next asked whether tumor mutation burden (TMB) or likely pathogenic somatic mutation burden (LPTMB) differed across the ecDNA, chromosomally amplified, or nonamplified tumors. To test this, we fitted logistic regressions on tumor type, age at diagnosis, sex and batch (SJ or CBTN). No independent effect of TMB (Wald test; *p* = 0.74; *n* = 1749) or LPTMB (*p* = 0.51) was observed in amplified vs. nonamplified tumors (**Supplementary Figure 5c**); however, within the amplified tumor subset, ecDNA(+) tumors had lower LPTMB than chromosomally amplified (*p* = 0.007; *n* = 232), accounting for independent effects of the above covariates (**Supplementary Figure 5d**). These results indicate that age and tumor type are likely the strongest determinants of overall tumor mutation burden, but that tumors with extrachromosomal amplification have significantly lower LPTMB than tumors with chromosomal amplifications only.

### ecDNA is associated with poor survival in pediatric cancers

To investigate whether ecDNA in pediatric cancers is associated with worse clinical outcome, we performed Kaplan-Meier regression on overall survival data available for 1820 patients (**Supplementary Table 1**). Here, we stratified patients into those whose tumors had ecDNA (*n* = 216), chromosomal amplification only (*n* = 225), or no focal amplification (*n* = 1452). Kaplan-Meier regression indicated that patients with ecDNA(+) tumors had significantly worse outcomes than patients with tumors containing no amplification (log-rank test, adjusted *p* < 2e-16) and patients with chromosomal amplifications (adjusted *p* = 7e-6) (**Figure 4a**). To estimate the magnitude of the association between ecDNA and survival, we performed Cox regression of sex, age at diagnosis, tumor type, and amplification type (ecDNA, chromosomal, or no amplification). Controlling for available covariates, ecDNA detected in a patient tumor conferred twofold risk of death within 5 years relative to tumors without amplification (hazard ratio 2.1, *p* = 5e-9, Wald test), and one-and-a-half-fold risk relative to tumors with chromosomal amplification but no ecDNA (HR 1.5, *p* = 0.02, **Figure 4b**).

**Figure 4:**
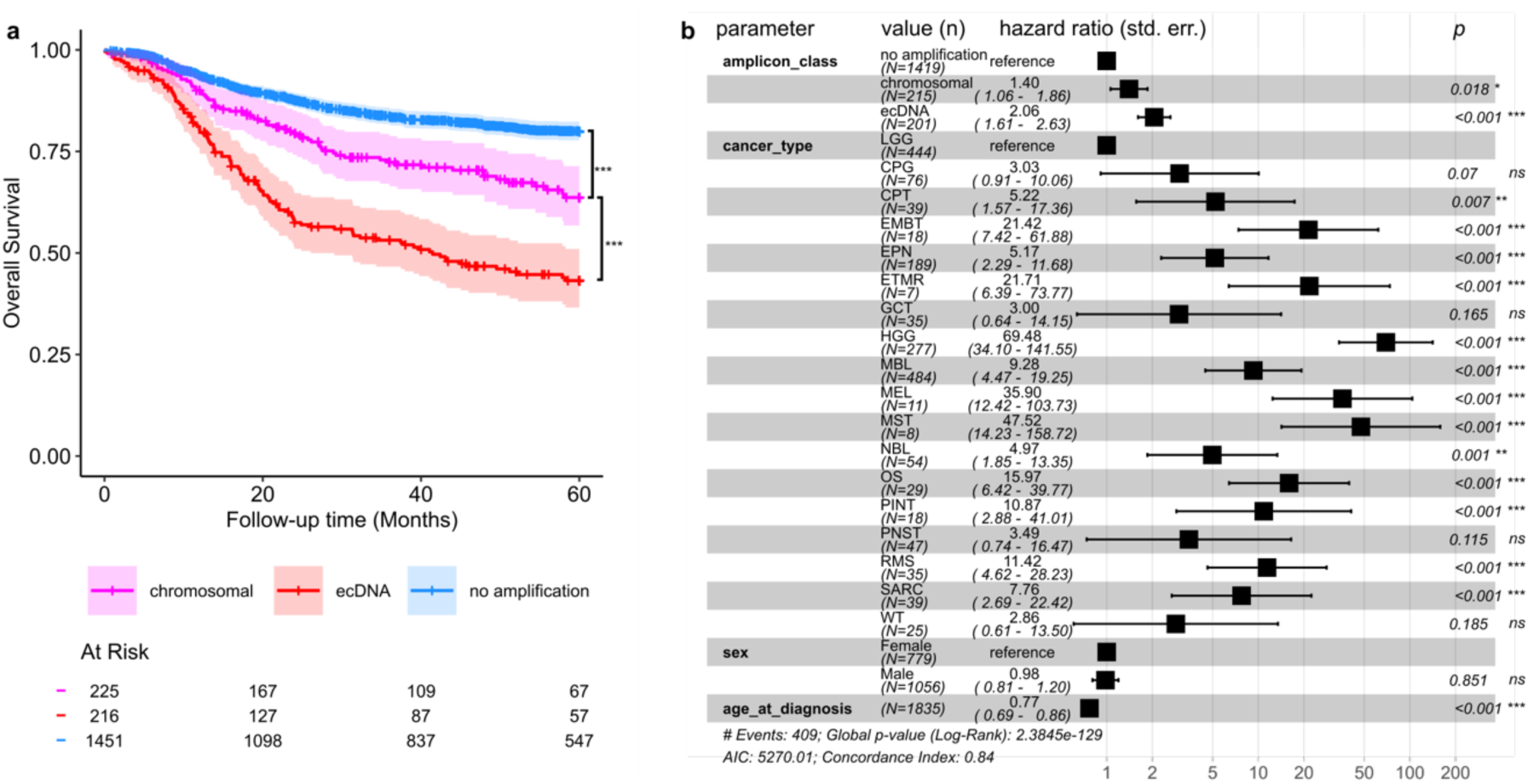
Overall survival of pediatric patients with ecDNA(+) tumors. Sample was limited to tumor types with at least 5 cases, at least 1 ecDNA(+) sample, and at least 1 clinical event. (**a**) Patients with ecDNA(+) tumors have significantly worse outcomes than patients with tumors containing no amplification (adjusted *p* < 2e-16) or patients with chromosomal amplifications (adjusted *p* = 7e-6; log-rank test with Benjamini-Hochberg correction). (**b**) Cox regression indicates twofold risk of death for patients with ecDNA compared those without amplifications (hazard ratio 2.1, *n* = 1836, *p* = 5e-9) and one-and-a-half-fold relative to patients with chromosomal amplifications only (HR 1.5, *p* = 0.02). ***𝑞<0.001; **𝑞<0.01; *𝑞<0.05; ns, 𝑞≥0.05.

Because childhood cancers are rare and highly diverse, survival analyses could not be performed robustly for every tumor type due to limited patient numbers within each category. To address this limitation, we integrated our cohort with previously published whole genome sequencing datasets from studies of medulloblastoma^7^, neuroblastoma^10^, and pediatric high-grade glioma^54^. This approach enabled us to investigate more thoroughly the association between ecDNA and patient outcomes within each of these three tumor types.

Joint modelling of focal amplification, ecDNA, molecular subgroup, and other clinical variables across 445 MBL tumors suggests that ecDNA (HR 2.8, *p* = 6e-4), rather than focal amplification of any kind (HR 0.9, *p* = 0.71) is a stronger predictor of 5-year survival (**Supplementary Figure 6a-b**). Similarly, survival regressions across 175 NBL tumors estimate poorer 5-year overall survival for ecDNA(+) tumors accounting for chromosomal amplifications (KM: *p* = 0.042, BH-adjusted log-rank test, **Supplementary Figure 6c**; Cox: HR = 2.8, *p* = 0.048, **Supplementary Figure 6d**), consistent with a previous report stratifying NBL tumors with focal amplification into high-risk with ecDNA amplification of *MYCN* vs. intermediate-risk with chromosomal amplification elsewhere in the genome^10^.

Myc family genes *MYC* and *MYCN* are major drivers of aggressive medulloblastoma^55^ and neuroblastoma^9,10^. We next investigated whether clinical outcomes may differ by the context of Myc family oncogene amplification, i.e. ecDNA or chromosomal. Across both cancer types, *MYC* and *MYCN* were amplified predominantly on ecDNA (MBL: 34/40, 85%; NBL: 27/31, 87%), leaving few chromosomally amplified cases as internal comparators. As expected, adjusting for Myc family gene amplification reduced the estimated hazard of ecDNA for both medulloblastoma (HR = 1.7, *p* = 0.14; **Supplementary Figure 7a-b**), and neuroblastoma (HR = 1.2, *p* = 0.78; **Supplementary Figure 7c-d**). We can therefore confirm that amplification of Myc family genes in these tumors in their predominantly extrachromosomal form carries an exceptionally poor prognosis, consistent with previous reports^10,55^; although the scarcity of chromosomally amplified *MYC*(*N*) tumors limits assessment of whether extrachromosomal Myc family amplification exerts further prognostic influence compared to chromosomal.

Molecular determinants of prognosis in pediatric high-grade gliomas include pathogenic mutations of histone 3 (H3), *IDH*, and *TP53* genes as well as focal amplification of diverse oncogenes. It has been previously shown that complex structural variants are associated with poorer overall survival of pHGG patients, where *TP53* loss and ecDNA were enriched in the complex-SV group^54^. Combining that cohort with pHGG tumors from our pancancer cohort, we confirmed that the strongest predictor of outcome was H3K27M mutation, which distinguishes high-risk diffuse midline gliomas (DMG) with universally poor prognoses (**Supplementary Figure 8a-b**). Further stratification by amplification status showed that the presence of chromosomal or extrachromosomal amplification was nominally but non-significantly associated with poorer survival within 5 years of diagnosis (**Supplementary Figure 8c**). Given the exceptionally poor outcomes associated with H3K27 alteration, we hypothesized that the presence of H3K27M mutation may supersede the prognostic utility of ecDNA in this tumor subtype. In a revised hierarchical model stratified by H3K27 alteration, we estimated that ecDNA is a prognostic indicator specifically in H3K27 wild-type tumors (HR = 1.81; *p* = 0.02); however, the estimated additional hazard of ecDNA in H3K27-altered tumors was nonsignificant and near-zero (HR = 1.2; *p* = 0.58; **Supplementary Figure 8d**). These findings indicate a context-dependent role for ecDNA in pHGG prognosis, where it is predictive of worse overall survival except in H3K27-altered tumors.

### ecDNA evolves during progression and recurrence of pediatric cancers

Recognizing that ecDNA may contribute to tumor evolution and treatment resistance, we next compared its prevalence in primary versus relapsed tumors and tracked its dynamics in longitudinal primary-relapse pairs.

For this analysis, we aggregated progression, recurrence, relapse, and metastasis into one category of secondary samples. Secondary malignancies and autopsy samples were excluded, as were tumor types without focal amplification. ecDNA was modestly but significantly more frequent in secondary (57 of 519, 11%) than primary tumors (202 of 2448, 8%), controlling for tumor type (**Figure 5a**; stratified permutation test, *p* = 0.002). Tumors with chromosomal amplifications alone were not enriched in secondary compared to primary tumors (**Figure 5b**; *p* = 0.45).

**Figure 5:**
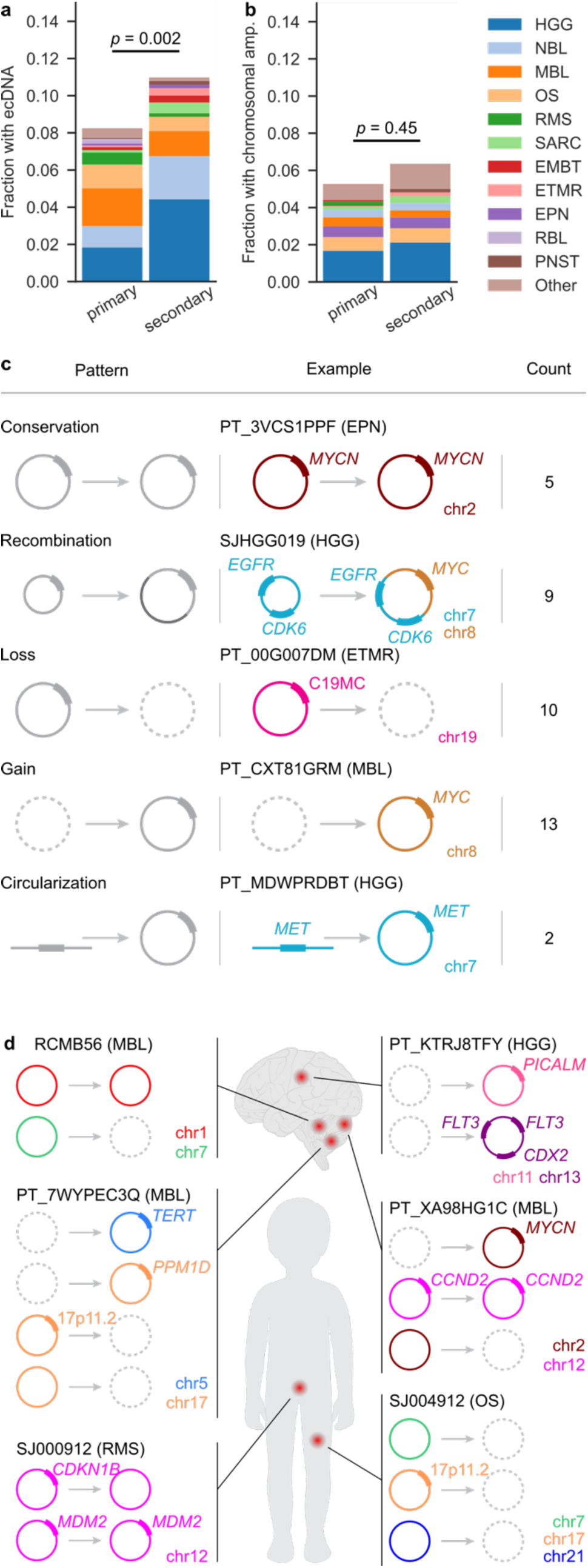
Sequence evolution of ecDNA during disease progression. (**a**-**b**) Proportion of (**a**) tumors with ecDNA and (**b**) tumors with chromosomal amplification only, by tumor type. *p*-values derived from permutation tests stratified by tumor type. (**c**) 39 ecDNA sequences were observed longitudinally in 29 tumors. We classified these based on ecDNA sequence over time: conservation (no change), recombination (sequence addition or loss to an extant ecDNA), loss (no amplification observed in the latter sample), gain (no amplification in the earlier sample), or circularization (chromosomal amplification at the same locus in the earlier sample). No examples were observed of linearization (chromosomal amplification at the same locus in the later sample). (**d**) Multiple ecDNA sequences were observed longitudinally in 6 tumors. Amplified oncogenes are annotated. EPN, ependymoma; ETMR, embryonal tumor with multilayered rosettes; HGG, high-grade glioma; MBL, medulloblastoma; RMS, rhabdomyosarcoma; OS, osteosarcoma.

To characterize patterns of ecDNA evolution, we collated 29 longitudinal cases where ecDNA was detected in at least one of sequential tumor samples of the same patient (**Supplementary Table 10**). Here, we included an ecDNA(+) medulloblastoma patient tumor diagnosed at Rady Children’s Hospital, San Diego^7^ and for which we generated new WGS data from the relapse. In these 29 longitudinal cases, we observed 39 distinct ecDNA sequences. Of these, 13 ecDNA arose *de novo* compared to earlier tumor samples (gain) and 10 ecDNA were not detected in a later tumor sample (loss). Two chromosomal amplifications were identified as ecDNA in a later tumor sample (circularization). We do not observe examples of ecDNA amplification later reclassified as non-ecDNA chromosomal amplification. For extrachromosomal amplifications detected in the same patient at more than one timepoint, we asked whether the ecDNA had undergone structural variation in the interim, using a heuristic similarity metric incorporating Jaccard similarities of the amplified sequences and breakpoints (**Methods**). Five such ecDNA amplicons were conserved (*S* > 0.7) whereas the other nine showed structural evolution, including examples where a new oncogene was added (**Figure 5c**) or lost (**Supplementary Figure 9**) in the later tumor. For example, in a pHGG patient (SJHGG019), we observe the oncogenes *EGFR* and *CDK6*, both coded on chr7, co-amplified on the same ecDNA. The ecDNA observed in the relapse tumor of the same patient contained *MYC*, coded on chr8, as a third oncogene on the same ecDNA together with *EGFR* and *CDK6* (**Figure 5c**). Overall, these results suggest clonal tumor dynamics in which ecDNA may emerge, mutate, remain conserved, or be lost during disease progression.

Patient tumors with more than one ecDNA sequence showed longitudinal variability in the combinations of ecDNA sequences detected. For example, we previously observed two distinct ecDNAs, originating from chromosomes 1 and 7 respectively, present in a primary SHH medulloblastoma tumor^7^. In WGS data from the recurrent tumor of the same patient, we observe conservation of the ecDNA amplification originating from chromosome 1, whereas the ecDNA amplification originating from chromosome 7 was no longer detected in the relapse tumor. This and other examples (**Figure 5d**) highlight that multiple distinct ecDNA sequences provide tumors with flexibility to amplify different oncogenes and oncogene combinations during disease progression.

## DISCUSSION

By leveraging childhood cancer data cloud repositories with whole-genome sequencing data, we estimate frequencies of circular extrachromosomal amplification across a wide spectrum of childhood cancer types. Where grading of the tumor was incorporated into the histological or molecular diagnosis, the higher-grade diagnoses (pediatric high-grade glioma, choroid plexus carcinoma, malignant peripheral nerve sheath tumor, and pineoblastoma) had greater incidence of ecDNA than the lower-grade diagnosis (pediatric low-grade glioma, choroid plexus papilloma, neurofibroma, and low-grade pineal tumors), suggesting that ecDNA may be more frequent in pediatric tumors with high-grade histology across tissue origins. Notable high-grade tumor types in which we did not observe ecDNA are defined by well described oncogenic mutations, such as biallelic loss of *SMARCB1/SMARCA4* in atypical teratoid rhabdoid tumors (ATRT)^56^, *EWSR1*::*FLI1* (also commonly *EWS*/*FLI*) in Ewing sarcoma, or *ZFTA*^56^ gene fusions in supratentorial ependymoma. Consistent with a “Goldilocks principle”, excessive expression of *EWSR1*::*FLI1* is deleterious in Ewing sarcoma^57^, rendering high-copy amplification on ecDNA disadvantageous. A similar phenomenon may constrain *ZFTA* fusions in ependymoma, where ecDNA amplification would push fusion oncofactor activity beyond a tolerable range^58^. By contrast, solid tumor types where drivers benefit from gene dosage escalation more frequently contain ecDNA. Although we did not comprehensively analyze hematological cancers, a pediatric case report^59^ and a recent retrospective analysis of adult leukemias^60^ recommend this area for further investigation.

Patients with tumors containing ecDNA had significantly poorer survival compared to those with chromosomal amplification and those without amplification, controlling for sex, age and tumor type. This finding suggests that both focal amplification and extrachromosomal amplification have nonredundant prognostic value in pediatric cancers. The majority of ecDNA(+) tumors contained at least one well described oncogene, such as *MYC, MYCN*, *CDK4*, and others. In some tumors, multiple oncogenes were amplified on one or multiple ecDNA sequences. Other ecDNA amplifications contained genes not annotated in the COSMIC^16^ and ONGene^17^ oncogene databases but with plausible oncogenic roles, including *AKT3, NTN1,* and others. In rhabdomyosarcoma and retinoblastoma, we found recurrent gene fusions on ecDNA. The potential oncogenic roles of still many other ecDNA-amplified sequences without known oncogenes are less clear. For example, relatively little is known of the role of chr17p11.2 amplification, the most frequent ecDNA amplification in osteosarcomas. Intriguingly, this locus is a recombination hotspot in medulloblastoma^26^ and in some nonmalignant genetic disorders^61,62^. Given the current poor prognosis of osteosarcomas and the need for more effective treatment protocols, further analysis of this and other recurrently amplified loci are warranted to identify novel oncogenic drivers and treatment targets.

Across a set of paired primary/secondary cases, we observed various structural rearrangements of ecDNA sequences. This adds to accumulating evidence^2,7,63^ that ecDNA facilitates tumor evolution in response to therapeutic or other selective pressures. Secondary tumors contained ecDNA more frequently than primary tumors, controlling for tumor type, consistent with a previous analysis of adult tumors^63^. Due to limited treatment information available for this cohort, the role of extrachromosomal DNA evolution in the emergence of drug resistance under standard-of-care treatment including chemo- and radiotherapy remains to be further elucidated and functionally tested in pediatric solid tumor models.

Overall tumor mutation burden (TMB) was not associated with amplification or with ecDNA; however, ecDNA-amplified tumors had fewer somatic simple variants with predicted functional consequence targeting cancer genes than chromosomally amplified tumors. Speculatively, a possible explanation for this observation is that ecDNA may be sufficient to drive tumor growth with fewer secondary driver mutations. Focal amplification was specifically associated with somatic alterations to DNA damage response gene *TP53* and to chromosomal pairing and segregation components *KNL1*, *AURKA*, *STAG2*, and *KNSTRN*; however, no specific association was observed for ecDNA vs. chromosomal amplification. Should this observation prove generalizable, a possible explanation is that mechanisms for ecDNA generation associated with *TP53* inactivation such as chromothripsis^64^ may be at least as likely to generate chromosomal forms of focal amplification; or that other p53-independent mechanisms of ecDNA generation may yet be found. Future work will be required to substantiate and functionally validate the predictors of ecDNA and their potential underlying mechanisms.

Identification of ecDNA in patient tumors holds promise in prognostication and targeted treatment. Although the spectrum of known and putative oncogenic sequences on ecDNA is diverse and tissue-specific, there is some hope that patients with ecDNA(+) tumors may benefit from future therapies targeting a common vulnerability. For example, a recent study demonstrated that combination treatment of an ecDNA(+) gastric tumor using inhibitors against the ecDNA-amplified *FGFR* oncogene and a CHK1 inhibitor enhances transcription-replication conflict and prevents ecDNA-mediated acquired resistance that otherwise develops under FGFR inhibition alone^65^. Our results show that this approach may also be applicable to some high-risk pediatric tumors. For example, we recently reported preliminary clinical real-world experience using avapritinib, a next-generation tyrosine kinase inhibitor, in pediatric and young adult patients with recurrent/refractory PDGFRA-altered HGG^66^. Avapritinib was well tolerated and led to initial radiographic responses in 3 out of 7 cases. In one patient, *de novo EGFR* amplification was observed in a metastatic lesion that progressed during treatment, indicating oncogene switching as a possible resistance mechanism. A promising avenue for future investigation is whether combination therapy with an ecDNA-targeting treatment may prevent ecDNA-mediated resistance. ecDNA may also hold promise for detection via liquid biopsy as genes identified on ecDNA including *MYCN*^67^, *MYC*^68^, and *C19MC*^69^ are currently under investigation as liquid biomarkers.

Overall, this study highlights the spectrum of pediatric cancer patients with poor prognosis who may benefit from ecDNA-directed therapies. To facilitate future discovery, the dataset is available through interactive web portals at https://ccdi-ecdna.org/ and https://ampliconrepository.org/project/PedPanCan.

## Data Availability

WGS data from the PBTA and St. Jude datasets are under controlled access as implemented by the respective organizations but are available from the following sources upon approval from an institutional data access committee. PBTA patient cohort: Kids First Data Resource Center (https://kidsfirstdrc.org) via the CAVATICA data portal. Inclusion criteria were patient solid tumors with WGS in the OpenPBTA, X01, and PNOC datasets. St. Jude patient cohort: St. Jude Cloud (https://www.stjude.cloud). Inclusion criteria were patient solid tumors with WGS from the Pediatric Cancer Genome Project (PCGP; SJC-DS-1001), Clinical Pilot (SJC-DS-1003); Genomes 4 Kids (G4K; SJC-DS-1004), Real-Time Clinical Genomics (RTCG; SJC-DS-1007); and Pediatric Brain Tumor Portal (PBTP; SJC-DS-1014) datasets as of September 2022. WGS data for additional ETMR and spinal ependymomas will be made available upon reasonable request.
All source metadata are available in the Supplementary Tables and/or from the cited sources. Data and derived visualizations describing all amplifications are accessible from the data portals at https://ccdi-ecdna.org/ and https://ampliconrepository.org/project/PedPanCan.

https://ampliconrepository.org/project/PedPanCan

## SUPPLEMENTARY FIGURES

**Supplementary Figure 1:**
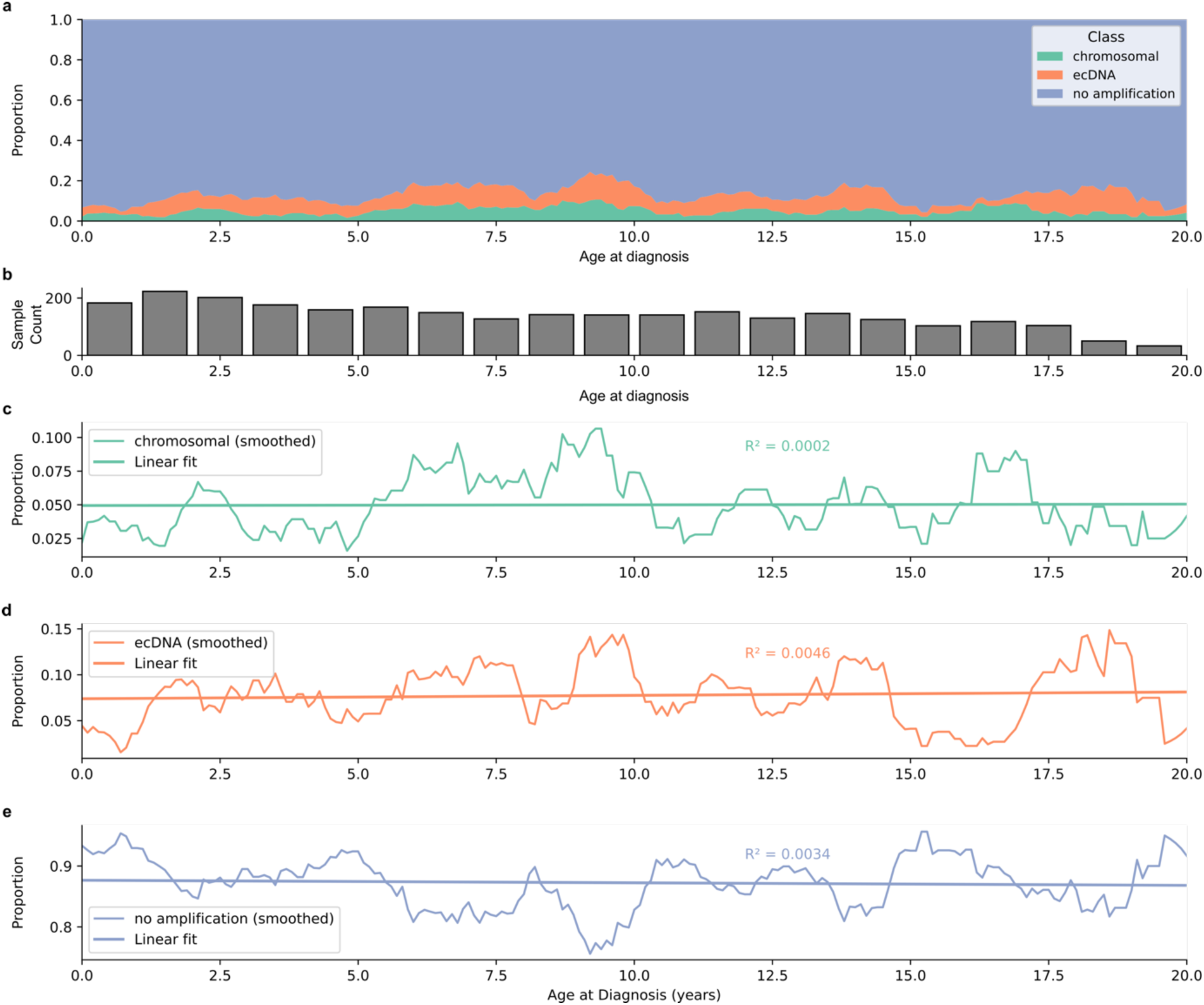
Ordinary least squares regression of focal amplifications in patient tumors on age at diagnosis. (**a**) Ages at diagnosis were binned to 0.1-year resolution, and class proportions (ecDNA, chromosomal amplification, or no amplification) were computed for each age group. To reduce sampling noise, a centered 1-year rolling average was applied to the age-wise amplicon class distributions. (**b**) Distribution of sample counts by age. (**c**-**e**) Ordinary least squares linear regressions for proportion of tumors with chromosomal amplification (𝑅^2^= 2e − 4, 𝑝 = 0.84, Student’s *t*-test), ecDNA (𝑅^2^= 0.005, 𝑝 = 0.38), or no amplification (𝑅^2^= 0.003, 𝑝 = 0.41).

**Supplementary Figure 2:**
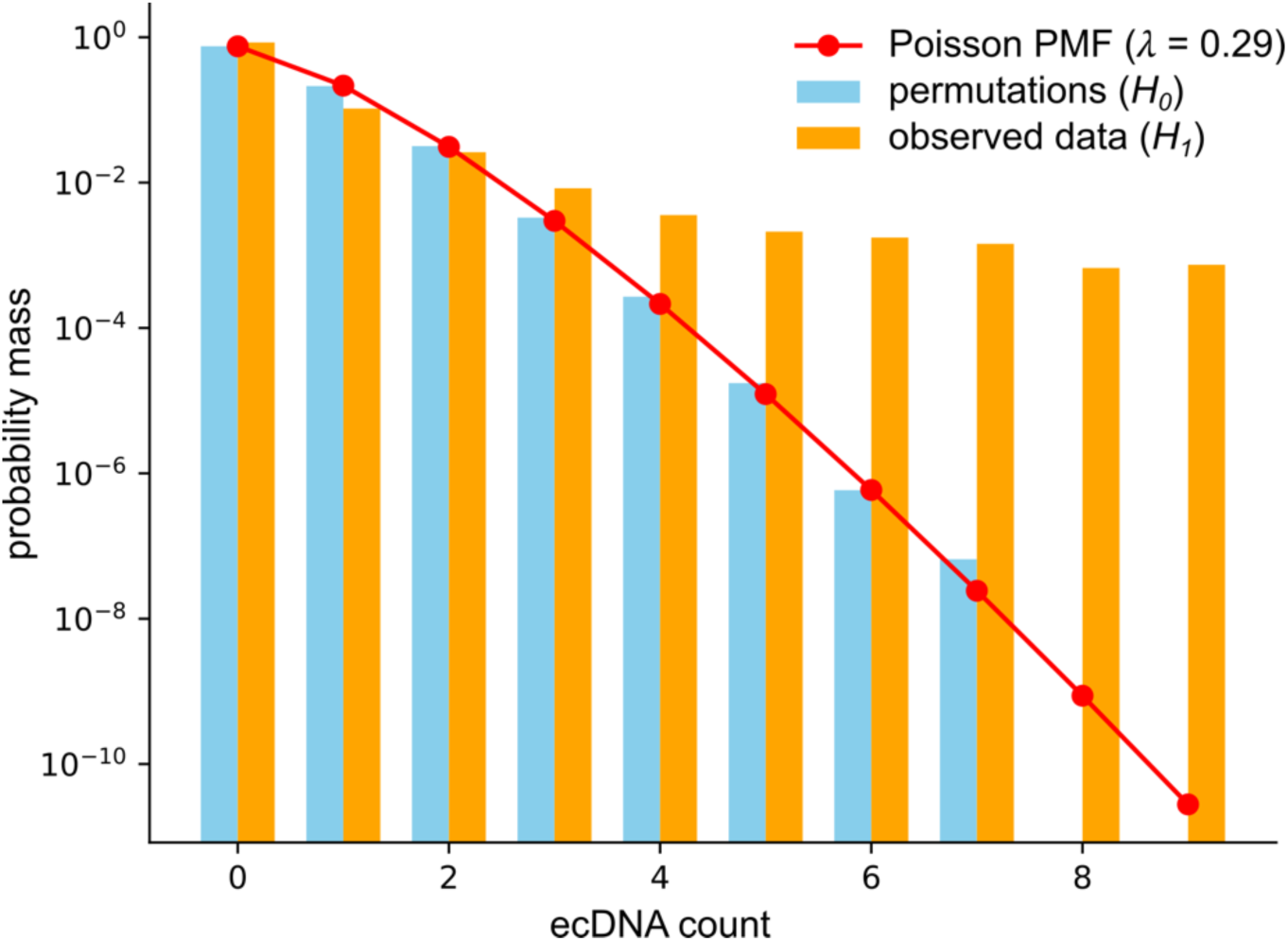
Expected and observed distributions of ecDNA across the human reference genome. A null hypothesis *H_0_* in which ecDNA is uniformly and randomly distributed across the genome was modelled explicitly as a Poisson distribution with rate parameter 𝜆 proportional to the number and average length of ecDNA sequences (red circles) and experimentally by shuffling the locations of ecDNA across the genome (blue bars). 99.9% of the probability mass of *H_0_* falls within the range 0 ≤ n < 3, such that the chosen threshold n ≥ 3 corresponds to a p-value threshold of *p*=0.001. By contrast, the observed distribution of ecDNA (*H_1_*, orange bars) was concentrated at some genomic regions, indicating a nonrandom distribution of ecDNA in pediatric cancer genomes. PMF, probability mass function.

**Supplementary Figure 3:**
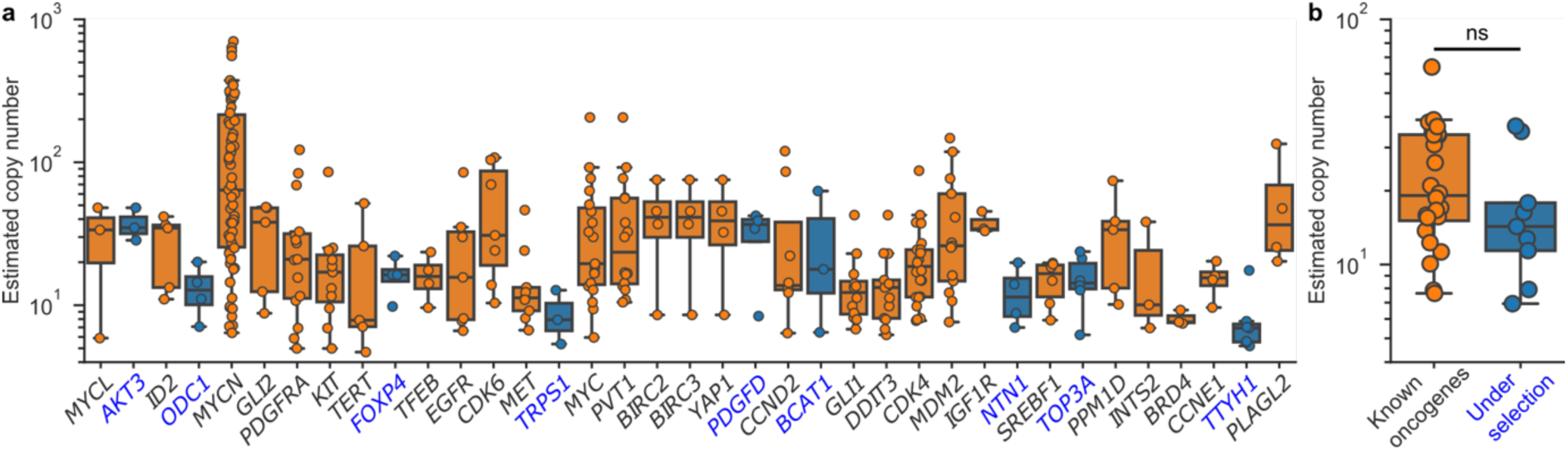
Copy number of genes recurrentlu amplified on ecDNA. (**a**) Estimated copy number of each amplification for genes of interest indicated in Figure 2. (**b**) Medians of gene copy numbers in (**a**); two-sided Mann-Whitney *U* test; 𝑈 = 138; 𝑝 = 0.23. Medians of *PVT1*, *BIRC2*, *BIRC3* and *DDIT3* were excluded for violations of the indpendence assumption.

**Supplementary Figure 4:**
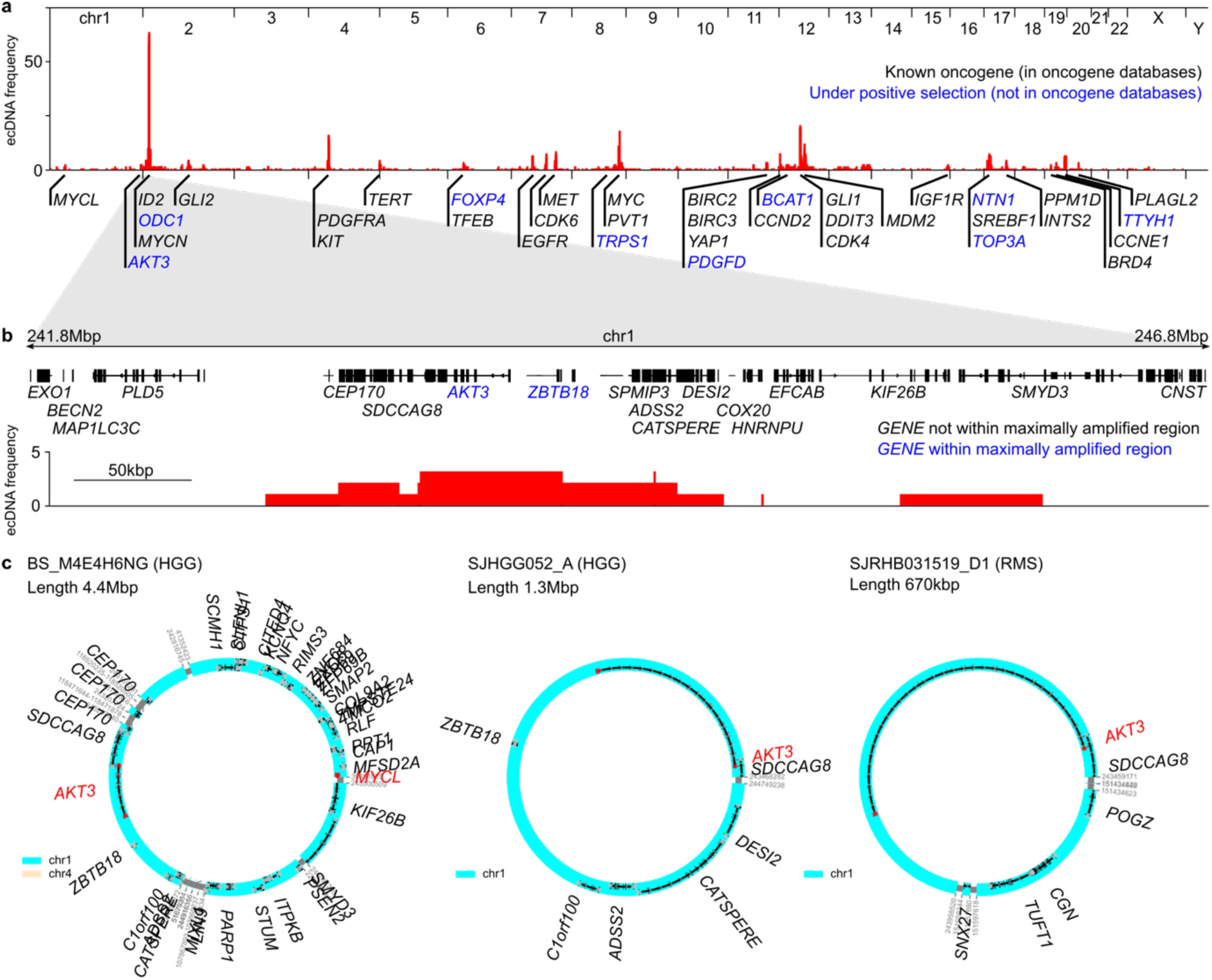
A recurrently ecDNA-amplified locus contains putative oncogene *AKT3*. (**a**) Panel reproduced from Fig. 2a. Chromosomal co-ordinates of ecDNA amplifications were stacked to generate a histogram of ecDNA amplifications across the genome. Selected recurrently amplified (𝑛 ≥ 3) genes of interest are highlighted. (**b**) Histogram data from (a) zoomed in to show a 5Mbp peritelomeric portion of chr1 containing the *AKT3* locus. The consensus genomic region amplified in all 3 tumors, or maximally amplified region, contains 2 complete protein-coding gene loci: *AKT3* and *ZBTB18*. (**c**) Circular sequence assemblies of the ecDNA sequences containing the full-length *AKT3* locus. Known and putative oncogenes *AKT3* and *MYCL* are highlighted in red.

**Supplementary Figure 5:**
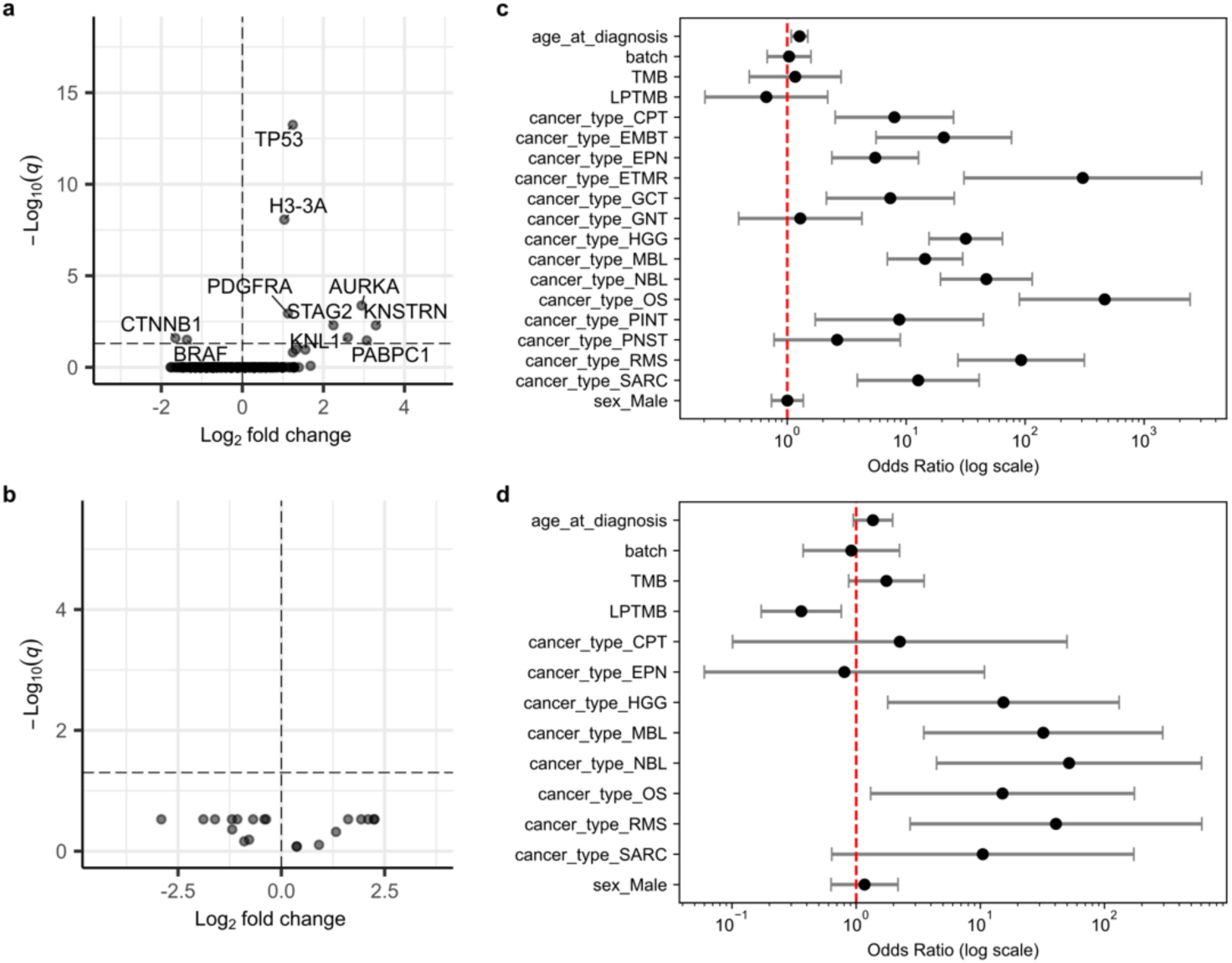
Tumor mutation burden of ecDNA(+) and chromosomally amplified tumors. (**a**-**b**) Associations of “likely pathogenic” somatic simple variants, defined as simple variants with predicted functional consequence targeting cancer-related genes, in (**a**) amplified vs. nonamplified and (**b**) ecDNA(+) vs. chromosomally amplified pediatric tumors (*n* = 1798). Genes are labelled in which pass the *q* < 0.10 significance threshold (*χ^2^* test with BH correction) indicated by the horizontal bar. (**c**-**d**) Maximum likelihood logistic regression models of (**c**) amplified vs. nonamplified (*n* = 1749) and (**d**) ecDNA vs. chromosomal amplification (*n* = 232) on age at diagnosis, batch, TMB, LPTMB, tumor type and sex. Reference tumor type is low-grade glioma, LGG.

**Supplementary Figure 6:**
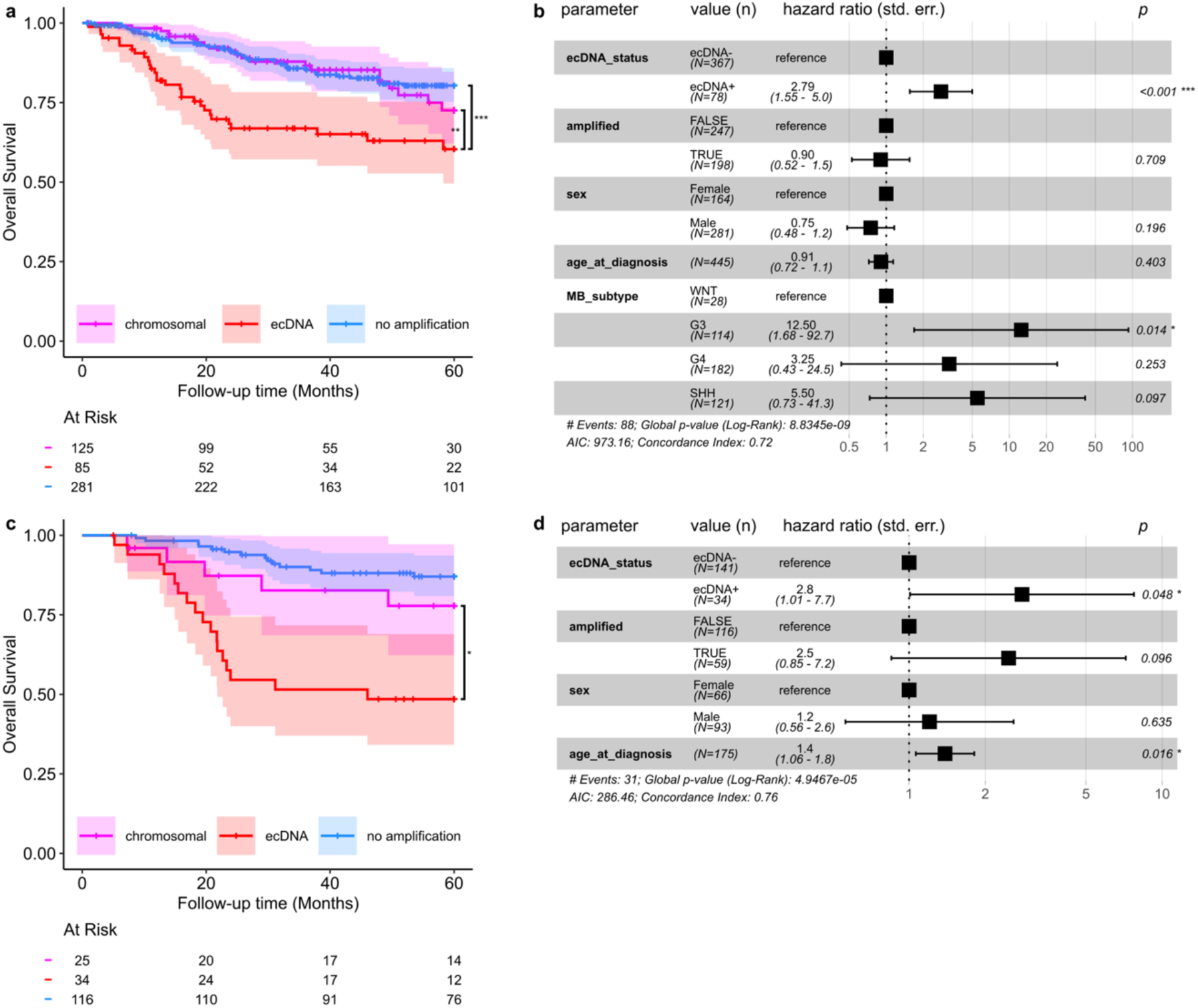
Survival of medulloblastoma and neuroblastoma tumors w.r.t. ecDNA. (**a**) Survival of 491 MBL tumors stratified by presence of chromosomal or extrachromosomal amplification. (**b**) Cox proportional hazards regression of 445 MBL tumors from (**a**) with complete metadata for molecular subgroup and clinical parameters. (**c**) Survival of 175 NBL tumors stratified by chromosomal or extrachromosomal amplification. (**d**) Cox proportional hazards regression of NBL tumors from (**c**) on amplification class and clinical parameters. * *p* < 0.05; ** *p* < 0.01; *** *p* < 0.001.

**Supplementary Figure 7:**
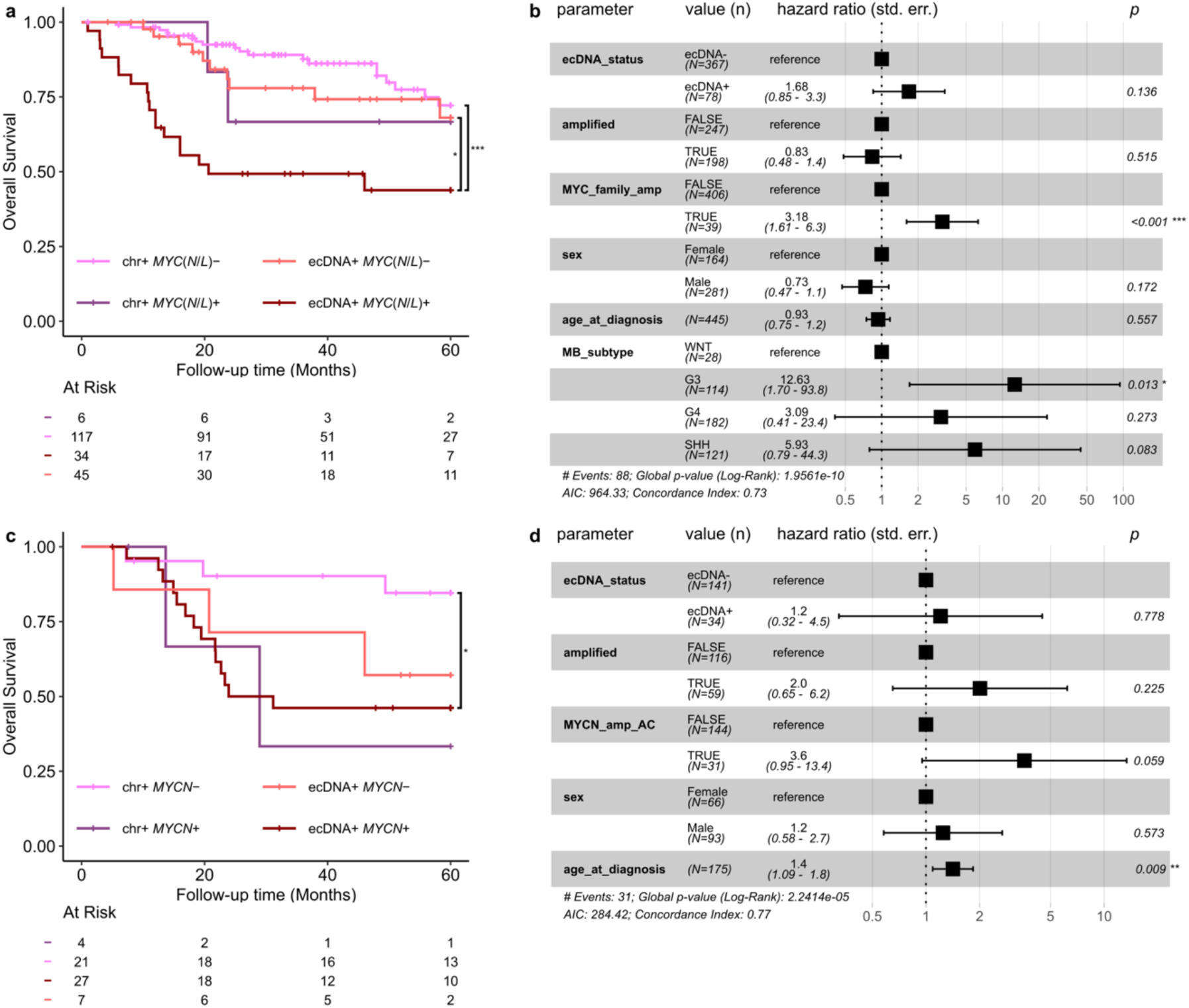
Survival of Myc family amplified medulloblastoma and neuroblastoma tumors. (**a**) Survival of 212 MBL tumors with focal amplification stratified by chromosomal/ecDNA and *MYC* gene family amplification. chr+, chromosomal amplification; *MYC*(*N*/*L*)+/−, *MYC* gene family amplified/nonamplified. (**b**) Cox regression as in (**Supplementary Figure 6b**), but including an additional covariate for *MYC* family amplification. (**c**) Survival of 59 NBL tumors with focal amplification, stratified by chromosomal/ecDNA and *MYCN* amplification. (**d**) Cox regression as in (**Supplementary Figure 6d**), but including an additional covariate for *MYCN* amplification. * *p* < 0.05; ** *p* < 0.01; *** *p* < 0.001.

**Supplementary Figure 8:**
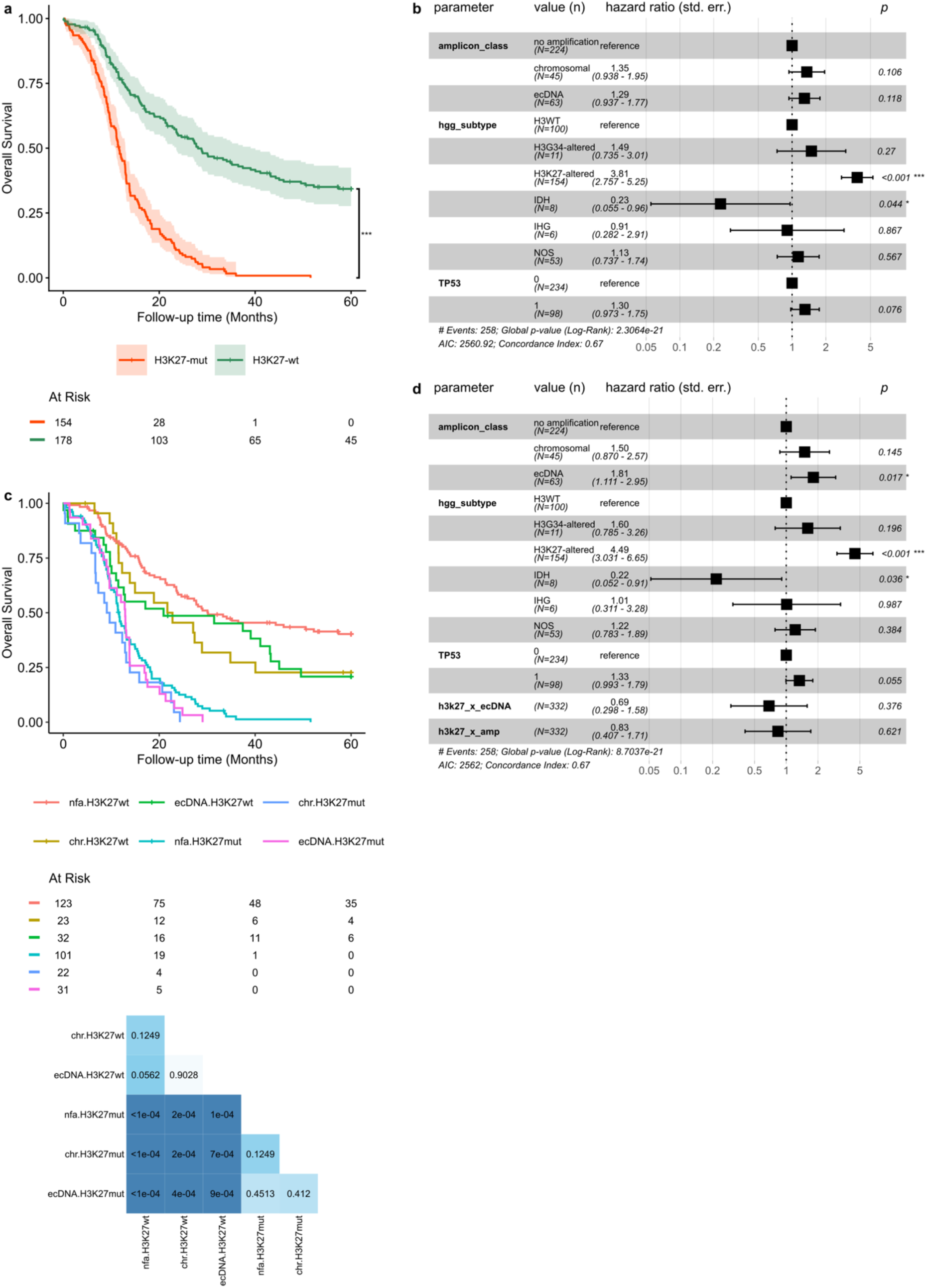
Survival of pediatric high-grade glioma tumors w.r.t. ecDNA. (**a**) Survival of 332 pHGG tumors stratified by chromosomal or extrachromosomal amplification. (**b**) Cox proportional hazards regression of pHGG tumors from (**a**) on amplification class and clinical parameters. (**c**) Survival of pHGG tumors as in (**a**), but stratified additionally on presence/absence of H3K27 alteration. nfa, no focal amplification; chr, chromosomally amplified; H3K27wt, H3K27 wild-type; H3K27mut, H3K27-altered. (**d**) Cox regression as in (**b**), but including additional interaction terms stratifying on amplicon class and H3K27 status. h3k27_x_ecDNA, H3K27 alteration and ecDNA; h3k27_x_amp, H3K27 alteration and focal amplification. * *p* < 0.05; ** *p* < 0.01; *** *p* < 0.001.

**Supplementary Figure 9:**
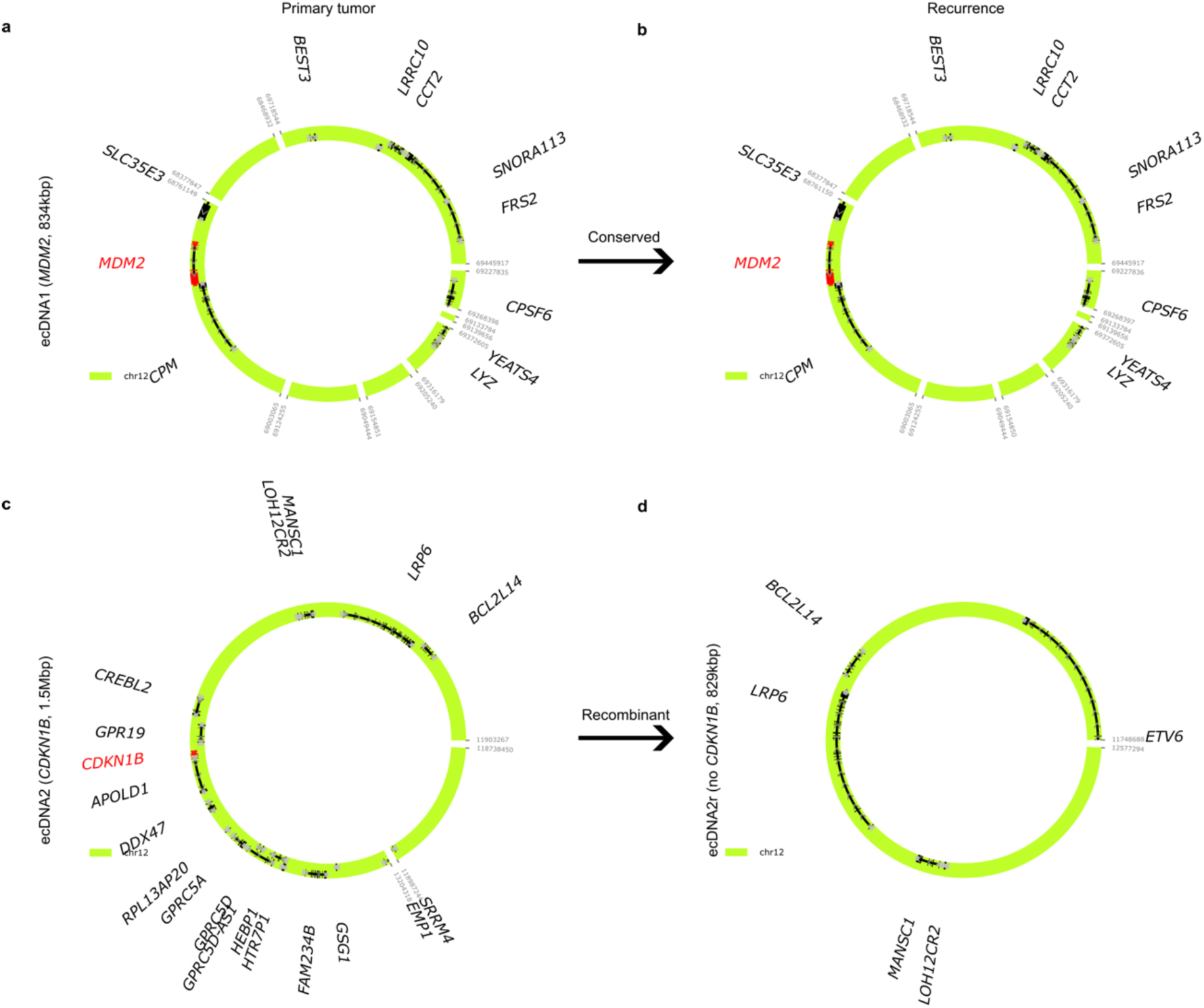
ecDNA amplifications detected in primary and recurrent tumors of rhabdomyosarcoma SJRHB012. (**a**-**b**) A conserved ecDNA sequence containing amplified *MDM2* which does not show evidence of sequence variation between (**a**) diagnosis and (**b**) recurrence biosamples of the rhabdomyosarcoma SJRHB012 (modified Jaccard similarity 0.75). (**c**-**d**) A nonconserved ecDNA sequence in the same tumor. The ecDNA sequence (**c**) in the primary tumor amplified a 1.5Mbp segment of chr12 including oncogene *CDKN1B*. The ecDNA sequence (**d**) in the same tumor at recurrence shared 674kbp of sequence homology with the primary ecDNA including *BCL2L14*, *LRP6*, *MANSC1*, and *LOH12CR2* but not *CDKN1B* (modified Jaccard similarity 0.21). All visualizations were generated using the cyclic sequence which explains the greatest proportion of copy number amplification.

## METHODS

### Pediatric tumor whole genome sequencing (WGS)

*Main cohort (PBTA+SJC).* Pediatric tumor WGS was identified from two pediatric cancer genomic data repositories, corresponding to 2397 biosamples from 1873 patients in the Pediatric Brain Tumor Atlas (PBTA) and Pediatric Neuro-Oncology Consortium (PNOC); and 1223 biosamples from 1094 patients in the St Jude Cloud (SJC)^12^ Pediatric Cancer Genome Project (PCGP; SJC-DS-1001)^70^, Clinical Pilot (SJC-DS-1003)^71^; Genomes for Kids (G4K; SJC-DS-1004)^72^, Real-Time Clinical Genomics (RTCG; SJC-DS-1007); and Pediatric Brain Tumor Portal (PBTP; SJC-DS-1014)^73^. Inclusion criteria were whole genome sequencing annotated with a solid tumor diagnosis; hematological malignancies, nontumor, and unannotated samples were excluded. Data preprocessing comprised each institution’s standard WGS bioinformatics pipelines. Alignments and genomic co-ordinates are w.r.t. human assembly GRCh38 (hg38).

### ecDNA detection and classification from bulk WGS

To detect ecDNA, samples in the WGS cohort were analyzed using AmpliconArchitect (AA)^1^ v1.2 and AmpliconClassifier (AC)^4^ v1.1.2. Briefly, the AmpliconArchitect algorithm was performed as follows. Copy number segmentation and estimation were performed using CNVkit v0.9.6^67^. Segments with copy number (CN) ≥ 4.5 were extracted as “seed” regions using AmpliconSuite-pipeline (April 2020 update)^74^. After confirming that sequencing depth was greater than 10x for all samples (mosdepth v0.3.8^75^), read coverage was downsampled to 10x. Then, for each seed, discordant read pairs indicative of genomic structural rearrangement were identified within and up to 50kbp distal the query region. Genomic segments were defined by genomic breakpoint locations (identified by discordant reads) and by modulations in genomic copy number. A CN-aware breakpoint graph of the amplicon region was constructed using the genome segments and breakpoints, and cyclic and non-cyclic paths are extracted from the graph. Amplicons were classified as ecDNA(+), breakage-fusion-bridge (BFB), complex non-cyclic, linear, or no focal amplification using the heuristic-based companion script AmpliconClassifier. Biosamples with one or more amplicons classified as ecDNA were labelled ecDNA(+). Biosamples without ecDNA but with one or more amplicons classified as BFB, complex non-cyclic or linear were labelled as chromosomally amplified. All other samples were annotated as having no focal amplifications.

All amplifications classified as ecDNA were manually reviewed for possible false positives, marking for reanalysis amplicons satisfying any of the following criteria:

- amplicon has high variability in depth of coverage of mapped reads, indicating a genomic locus of low mappability;
- cyclic subsequences which were assigned CN < 4.

These samples were reanalyzed using the latest AmpliconSuite versions as of Aug. 2024 (AA v1.4.r2, AC v1.2.0). Samples reclassified in this way were BS_2F8PFTW5, BS_XB34VS6P, BS_G6KYSGQF, BS_ GBSSZBMF, and BS_YMYESCY7.

### Patient metadata and tumor type annotations

Clinical metadata including sex, age at diagnosis, histology, and tumor history (primary, secondary, etc.) were available from the respective cloud genomics data platforms https://pedcbioportal.kidsfirstdrc.org/ (PBTA) and https://pecan.stjude.cloud/ (SJC), and from previous peer-reviewed publications^11,54^. Molecular diagnoses were integrated with histological diagnoses for biosamples included in the Pediatric Brain Tumor Atlas (PBTA)^11^ in the following priority order: molecular subtype, DKFZ v12 methylation classification (confidence threshold 0.9), DKFZ v11 methylation classification^13^ (likewise), harmonized diagnosis, original histological diagnosis. Diagnoses were subsequently collated and harmonized between the data sources to annotate the tumor types and subtypes described herein (**Supplementary Table 11**). Mutation status of high-grade gliomas for *TP53*, *IDH*, and H3 genes were extracted from diagnosis annotations.

Annotations of tumor purity were available for 2064 biosamples representing 1679 tumors from the PBTA cohort from a previous publication^11^ using the THetA2 software^76^. To annotate SJC samples, we applied THetA2 to an additional 1231 samples representing 1100 tumors.

### Supplementary tumor WGS

#### Additional MYCN-amplified spinal ependymomas

WGS data from 3 pediatric *MYCN*-amplified spinal ependymomas was peprocessed, and aligned according to best practices to human reference GRCh38. Focal amplifications and ecDNA were analyzed as above using AmpliconArchitect v1.5.r2 and AmpliconClassifier v1.3.3. Results were used to validate ecDNA amplification of *MYCN* in spinal ependymoma and were excluded from other analyses.

#### Additional ETMR tumors

WGS data of 38 biosamples from 30 ETMR tumors including 29 samples from a previous publication^27^ were uniformly reprocessed and aligned to human reference GRCh37. Amplicon reconstruction and classification was performed using default paramaterizations for AmpliconArchitect v1.5.r2 and AmpliconClassifier v1.3.7. To improve detection sensitivity of low-copy *C19MC* amplifications in 11 samples, AmpliconArchitect was re-run skipping the default copy number variant detection step and instead specifying the genomic co-ordinates of suspected *C19MC* amplifications according to the following rules: for samples where *C19MC* amplification was detected a different sample from the same tumor, the boundaries of that amplicon were used; and for tumors for which a low-copy gain was identified (copy number estimate from bulk WGS between 2.5 and 4.5), the boundaries of the low-copy gain were used. Results were used to validate ecDNA amplification of *C19MC* in ETMR and excluded from other analyses.

#### Additional pediatric high-grade gliomas

Previously published WGS data corresponding to 53 pediatric HGG tumors with accompanying clinical metadata including outcomes were accessed from the archival sequencing data repository^54^. Amplicon detection and classification was performed as in the main cohort above. Data were included in pHGG survival analyses and excluded elsewhere.

#### Additional neuroblastomas

Previously published amplicon annotatations and associated clinical metadata for 114 neuroblastoma tumors^10^ were included in survival analyses of NBL.

#### Additional medulloblastomas

Previously published amplicon annotatations and associated clinical metadata for 223 additional medulloblastoma tumors^7^ were also included in survival analyses and excluded elsewhere.

### Somatic variant data collection and analysis

#### Somatic variants

Consensus simple somatic variant data were retrieved from the respective institutional somatic variant calling and annotation pipelines which passed standard quality control thresholds^11,12^. Annotations were sourced from Variant Effect Predictor (VEP)^77^ and Ensembl release v105 (CBTN) or v100 (St Jude). Simple somatic variants were defined as likely pathogenic which satisfied the following criteria:

- affecting a known cancer gene, defined as tumor suppressor and oncogenes included in the COSMIC Cancer Gene Census v101;
- Ensembl consequence (IMPACT) HIGH or MODERATE;
- PolyPhen^78^ predicted functionality ‘possibly damaging’ or ‘probably damaging’ if available; and
- SIFT^79^ predicted functionality ‘deleterious’ or ‘deleterious_low_confidence’, if available.

For association testing, likely pathogenic somatic variants were aggregated at the gene level; ie., biosamples were considered altered at gene *G* which had any likely pathogenic variant mapping to *G*. Association was tested using the *χ^2^*test for independence with Bonferroni correction where at least 5 samples were altered at a given gene and at least 5 amplified; otherwise the association was considered underpowered and therefore not tested.

#### Germline variants

Annotations of pathogenicity for germline variants of the OpenPBTA subcohort were retreived from a previous publication^80^. Association testing w.r.t. focal amplification and ecDNA was performed as described above for somatic variants. Variant enrichment was plotted using EnhancedVolcano v1.24.0.

### Logistic regressions

For all logistic regressions, tumor types were included which had at least 5 cases and at least 1 ecDNA(+) and 1 chromosomally amplified case. Numerical covariates were z-score normalized according to best practice.

#### Tumor purity on amplicon class

To determine whether tumor purity influences ecDNA detection efficiency, we fitted logistic regressions of amplicon class (ecDNA, chromosomal amplification, or no amplification) on sex, age at diagnosis, and cancer type, including and excluding tumor purity (as estimated by ThetA2) as an additional covariate. Logistic regression models were implemented using scikit-learn v1.5.2^81^ using the Newton conjugate gradient solver and no penalty term. Models with and without tumor purity as an additional covariate were compared by likelihood ratio test.

#### Tumor mutation burden on amplicon class

Regressions were fitted on age at diagnosis, sex, source (CBTN or St Jude), tumor mutation burden (TMB), likely pathogenic mutation burden (LPTMB) and tumor type using statsmodels v0.14.6 and plotted using base matplotlib v3.10.3.

### Survival analyses

Patient overall survival data were included in PBTA clinical metadata and provided upon request from St. Jude. 223 medulloblastoma tumors from the International Cancer Genome Consortium and analyzed in a previous publication^7^ were also included in survival analyses. St. Jude survival data did not include date of most recent follow-up, so survival data were censored at the date of data collection. Tumors with linear, complex noncyclic, or breakage-fusion-bridge amplifications but without ecDNA were classified as chromosomal. Kaplan-Meier and Cox Proportional Hazards regressions were performed in R 4.5.3 with the survival^82^ v3.8.6 and survminer v0.5.2 packages.

#### Kaplan-Meier (KM) regressions

Sample was limited to tumor types with at least 5 cases, at least 1 death and at least 1 ecDNA(+) sample (*n* = 1892; 225 ecDNA(+), 216 with chromosomal amplification only, and 1451 unamplified). Differential survival reported for KM regressions was evaluated by log-rank test.

#### Cox Proportional Hazards (CPH) regression

Hazard ratios were estimated by CPH regression parameterized on amplification class, tumor type, sex and age at diagnosis, and evaluated by log-rank test. Sample was as in KM above but additionally required complete annotations for sex and age at diagnosis, resulting in 56 exclusions (*n* = 1836). Applicability of the proportional hazards assumption was validated by likelihood ratio test (𝛼 > 0.1) implemented in the ‘cox.zph’ function of the survival package.

### Ordinary least-squares linear regression

For regression of amplification status (ecDNA, chromosomal amplification, or no amplification) on age at diagnosis, ages were binned at 0.1-year resolution and a centered 1-year rolling average (smoothening) was applied to reduce sampling noise. For tumors with sampled more than once, the earliest biosample was used for determining amplification status. Ordinary least-squares linear regression models were then fit independently for each class using age as the predictor and smoothened class proportion as the response using scikit-learn^81^ v1.5.2. Significance testing was performed using Student’s *t*-test implemented in statsmodels^83^ v0.14.2.

### Permutation testing of amplification in primary and secondary tumors

To test whether ecDNA (or chromosomal amplification) is more frequent in secondary than primary tumors, a permutation test was constructed as follows. Let the test statistic 𝑇 = 𝑓_s_ −𝑓_p_, where *f_s_* and *f_p_* are the observed frequencies of ecDNA+ tumors among secondary and primary tumors respectively. Then, permutations are constructed as follows. For paired samples, (ie, the same tumor was sampled at diagnosis and again at progression), the primary/secondary labels are swapped with 50% probability. For unpaired samples, for each tumor type, primary/secondary labels are shuffled among samples of that tumor type. The permutation procedure thus samples from a null distribution for which the distribution of ecDNA across tumor types is unchanged, but without any association with primary/secondary status. 1000 permutations were performed and a *p*-value calculated according to standard practice.

### Other statistical methods

Statistical test, test statistic and *p*-values are indicated where appropriate in the main text. Categorical associations were established using the chi-squared test of independence if *n* > 5 for all categories and the Fisher exact test otherwise, both implemented in scipy.stats^84^ v1.10.2. Multiple hypothesis correction was performed where appropriate using the Benjamini-Hochberg correction implemented in statsmodels^83^ v0.14.2 and reported as *q*-values or adjusted *p*-values. All statistical tests described herein were two-sided unless otherwise specified. For all tests performed on patient or biosample sets which require an independence assumption, samples from the same case were deduplicated according to the following priority order:

- any biosample classified ecDNA(+);
- and if any biosample with focal chromosomal amplification;
- the most recent biosample.

### Gene fusions

Fusion genes were identified by adjacency on the ecDNA sequence assemblies. For samples from the PBTA subcohort, fusion gene expression was confirmed from RNA-seq data as described in the source publication^11^.

### Longitudinal subcohort (Supplementary Figure 9; Supplementary Table 10)

Patient tumors were included in the set of longitudinal cases which satisfied the following criteria:

*- St Jude and PNOC cohorts.* For multiple tumors from the same patient, exactly one is annotated as “diagnosis”, and the others are labelled progressive, recurrent, or metastasis;
*- PBTA X00 and X01 cohorts.* Multiple tumors from the same patient for which the difference between the dates of diagnosis of the initial and the most recent biosample exceeds 30 days.

Similarity scores reported between amplicons were calculated using AmpliconClassifier feature_similarity.py v1.1.2. The formal definition and motivation of the similarity score is described previously^15^.

Circular sequences illustrated in **Supplementary Figures 4** & **9** reflect the highest-copy cyclic sequence detected using AmpliconArchitect containing the oncogene of interest. Figures were generated using CycleViz v0.2.1.

## ACKNOWLEDGMENTS

This work is supported by a generous endowment by the Clayes Foundation to the Research Center for Neuro-Oncology and Genomics within the Rady Children’s Institute for Genomic Medicine, a Hannah’s Heroes St. Baldrick’s Scholar Award (L.C.), the Dragon Master Foundation (L.C.), the Takeda Science Foundation (O.S.C.), funding from the National Institutes of Health (NIH) National Institute of Neurological Disorders and Stroke Institute R01NS132780 (L.C.), R21NS130137 (L.C.) and R21NS120075 (L.C.), the NIH National Cancer Institute P30CA030199 (L.C. and K.Y.), P30CA030199-42-S1 (L.C. and K.Y.), F31CA271777 (O.S.C.) U01CA253547 (J.P.M.), P30CA051008 (J.P.M.), U24CA220341 (J.P.M.), U24CA295532 (J.P.M.), and U24CA264379 (V.B. and J.P.M.), the NIH National Library of Medicine T15LM011271 (O.S.C.), the Japan Society for Promotion of Science (JSPS) 25K23852 (O.S.C.), 24KF0252 (O.S.C.), and Postdoctoral Research Fellowship F24404 (O.S.C. and D.K.), and a Moores Cancer Center Pilot Grant (L.C., V.B., and J.P.M.). This project is also supported by the UCSD/Rady Children’s Hospital pediatric hematology oncology fellowship program (S.S.). D.R.G. gratefully acknowledges support by the Clinician Scientist Program of the Mildred Scheel Cancer Career Center HaTriCS^4^ at the University Medical Center Hamburg-Eppendorf and the Else Kröner-Fresenius Stiftung (Grant 2025_EKEA.51). This work used Expanse cluster compute services at the San Diego Supercomputer Center through allocation BIO210026 from the Advanced Cyberinfrastructure Coordination Ecosystem: Services & Support (ACCESS) program, which is supported by National Science Foundation grants #2138259, #2138286, #2138307, #2137603, and #2138296. Support through grant P30CA030199 to the Genomics core facility at Sanford Burnham Prebys (NCI designated Cancer Center) is gratefully acknowledged. The content is solely the responsibility of the authors and does not necessarily represent the official views of the National Institutes of Health. This research was conducted using data made available by The Children’s Brain Tumor Network (formerly the Children’s Brain Tumor Tissue Consortium). This study makes use of data generated by the St. Jude Children’s Research Hospital – Washington University Pediatric Cancer Genome Project and Pediatric Brain Tumor Portal; and the St. Jude Children’s Research Hospital Genomes for Kids Study, Clinical Pilot and Real-Time Clinical Genomics projects. We thank J.H. Zhang, C. McLeod, D. Miller, Y. Zhu, D. Higgins and A. Resnick for facilitating data access, A. Calatroni for help with statistical analyses, R.J. Nowling for his open-source implementation of the likelihood ratio test, R. Wachs for figure design assistance, K. Ichimura and F.P.B. Dubois for scientific discussion, and anonymous reviewers for thoughtful and critical feedback.

## CONTRIBUTIONS

Conceptualization: S.S., O.S.C., J.P.M., and L.C. Methodology: O.S.C., S.S., J.L., J.P.M., and L.C. Investigation: O.S.C., S.S., E.Y.-C.C., R.K., J.K., A.D., S.Q.W., W.Z., H.H., and J.W. Formal analysis: O.S.C., S.S., E.Y.-C.C., R.K., R.G. Data curation: O.S.C., S.S., E.Y.-C.C., J.L., S.Z.W., E.R.-F., V.B., K.Y., J.P.M., and L.C. Software: O.S.C., S.S., E. Y.-C.C., R.K., J.K., A.D., S.W., W.Z., H.H., M.B., J.L., and Y.Y.L. Resources: J.L., K.O., D.R.G., J.G., D.K., K.Y., J.P.M., and L.C. Validation: Y.Y.L., E.R.-F., A.G.H., K.O., D.R.G., K.W.P., S.Z.W., J.G., and M.K. Visualization: O.S.C., S.S., and E.Y.-C.C. Writing – original draft: O.S.C. and S.S. Writing – review and editing: O.S.C., S.S., E.Y.-C.C., J.L., D.K., V.B., J.P.M., and L.C. Funding acquisition: O.S.C., D.K., V.B., K.Y., J.P.M., and L.C. Supervision: V.B., D.K., M.P., K.Y., J.P.M., and L.C. Project administration: L.C.

## RESOURCE AVAILABILITY

### LEAD CONTACT

Requests for further information and resources should be directed to and will be fulfilled by the lead contact, Lukas Chavez.

## MATERIALS AVAILABILITY

This study did not generate new unique reagents.

## DATA AND CODE AVAILABILITY

WGS data from the PBTA and St. Jude datasets are under controlled access as implemented by the respective organizations but are available from the following sources upon approval from an institutional data access committee. PBTA patient cohort: Kids First Data Resource Center (https://kidsfirstdrc.org) via the CAVATICA data portal. Inclusion criteria were patient solid tumors with WGS in the OpenPBTA, X01, and PNOC datasets. St. Jude patient cohort: St. Jude Cloud (https://www.stjude.cloud). Inclusion criteria were patient solid tumors with WGS from the Pediatric Cancer Genome Project (PCGP; SJC-DS-1001), Clinical Pilot (SJC-DS-1003); Genomes 4 Kids (G4K; SJC-DS-1004), Real-Time Clinical Genomics (RTCG; SJC-DS-1007); and Pediatric Brain Tumor Portal (PBTP; SJC-DS-1014) datasets as of September 2022. WGS data for additional ETMR and spinal ependymomas will be made available upon reasonable request.

All source metadata are available in the Supplementary Tables and/or from the cited sources. Data and derived visualizations describing all amplifications are accessible from the data portals at https://ccdi-ecdna.org/ and https://ampliconrepository.org/project/PedPanCan.

Source code for the AmpliconArchitect family of software tools is available at:

AmpliconSuite-pipeline: https://github.com/AmpliconSuite/AmpliconSuite-pipeline consisting of submodules:

- AmpliconArchitect: https://github.com/AmpliconSuite/AmpliconArchitect
- AmpliconClassifier: https://github.com/AmpliconSuite/AmpliconClassifier
- CycleViz: https://github.com/AmpliconSuite/CycleViz/.

Source code for software applets used to run AmpliconSuite-pipeline and other tools on the DNANexus (St. Jude Cloud) and CAVATICA (PBTA) platforms are available at:

- https://github.com/chavez-lab/ampliconsuite-dnanexus-applet
- https://github.com/chavez-lab/ampliconsuite-cwl-workflows
- https://github.com/jbk708/theta-applet

Source code for variant calling and annotation pipelines are available at:

- https://github.com/kids-first/kf-somatic-workflow
- https://github.com/kids-first/kf-annotation-tools

Source code to regenerate analyses and figures is available at https://github.com/auberginekenobi/pedpancan_ecdna.

