## Supplementary Notes for "Extrachromosomal DNA associates with poor survival across a broad spectrum of childhood solid tumors"

**Supplementary Note 1**

*Statistical basis for enrichment of ecDNA at a given genomic location.*

In assessing the distribution of ecDNA across the human genome, the question arises whether repeated observations of ecDNA at the same genomic location represents positive selection at that locus, or whether such observations could have occurred by chance. Here we show that a statistical threshold for this distinction is a function of the total base pair length of ecDNA observed in the dataset, which for this pediatric cohort corresponds to a threshold of *n* ≥ 3 independent cases with ecDNA at the same locus.

Assuming a null hypothesis *H_0_* where ecDNA sequences are independently and uniformly distributed across the mappable genome, the fraction of bases in the reference genome overlapping 1, 2, 3, etc. ecDNA sequences is expected to be Poisson-distributed, with rate parameter 𝜆 proportional to the number and average length of ecDNA sequences. To illustrate this, we sampled *H_0_* by randomly shuffling the locations of ecDNA sequences in our dataset and calculating the per-base coverage across the reference genome (**Supplementary Figure 2**). The sampled distribution (blue bars) closely matches the theoretical Poisson distribution (red circles); however the actual distribution of ecDNA across the genome (orange bars) has a heavier tail, suggesting positive selection at some genomic locations. 99.9% of the probability mass of *H_0_* falls within the range 0 ≤ *n* < 3, such that the chosen threshold *n* ≥ 3 corresponds to a p-value threshold of *p*=0.001 for significant enrichment of ecDNA at a given genomic region.

**Supplementary Note 2**

*Reanalysis of medulloblastoma tumors with respect to ecDNA*.

168 unique tumors previously analyzed using AmpliconArchitect^1^ were reanalyzed here, comprising a subset of that cohort from the PBTA and SJC datasets. Of these, 7 were reannotated with discrepant ecDNA classifications, and all were annotated previously as containing ecDNA (ecDNA+) but no ecDNA was detected in our reanalysis (ecDNA-). The discrepant ecDNA classifications are sufficient to explain the alternative estimates of ecDNA+ medulloblastomas as 18% in our previous publication and 16% herein.

In 1 of 7 examples, a low-confidence cyclic sequence was previously reported within a noncyclic focal amplification. No cyclic subsequence was detected in the reanalysis. As the evidence for ecDNA in this tumor is relatively weak, it is reclassified here.

In 6 of 7 examples, a low-copy (4 < *n* < 6) amplification was detected previously during the copy number calling step (CNVkit 0.9.7) which was determined to be a circular sequence assembly upon analysis with AmpliconArchitect. We have used similar methods in our reanalysis, but have altered the copy number calling step such that CNVkit runs in “paired” mode to estimate somatic copy number variation using whole genome sequencing (WGS) of tumor tissue and matched normal blood if available. No copy number variant was reported at these loci in the reanalysis, and therefore the examples were reclassified as ecDNA- in these 6 of 7 cases. We think it probable that the sensitivity of our Methods to classify ecDNA+ tumors is sensitive to the recall of low-copy focal somatic copy number amplifications (lcfCNVs) reported during the copy number calling step. Because a low-copy cyclic sequence assembly may indicate either circular extrachromosomal DNA or intrachromosomal tandem duplication (homogeneously staining region, HSR), we are unable to determine from WGS data alone whether these DNA sequences represent ecDNA or HSR.

No significant difference was observed in the fraction of medulloblastoma tumors classified as ecDNA+ between this cohort and the previous (*χ^2^* = 1.2, *p* = 0.55).

**Supplementary Note 3**

*CNS tumor types with infrequent incidence of ecDNA.*

*Ependymomas*

Among 239 ependymomas, 3 cases had ecDNA (1%). In one case, *MYCN* was amplified on ecDNA in primary and recurrent spinal tumors of the same patient. *MYCN*-amplified spinal ependymomas have recently emerged as an aggressive subtype of ependymoma^2^. In the remaining two cases, no further subtyping information was available. No known oncogenes were amplified on ecDNA, rendering these cases of unknown significance. Further subgrouping information for the other 236 ecDNA(-) ependymoma tumors was available as follows: posterior fossa group A (*n* = 50), posterior fossa group B (*n* = 9), posterior fossa myxopapillary (*n* = 2), posterior fossa not otherwise specified (NOS, *n* = 51), myxopapillary (*n* = 16), supratentorial *ZFTA* fusion (*n* = 24), supratentorial *YAP1* fusion (*n* = 2), supratentorial NOS (*n* = 15), non-*MYCN* spinal (*n* = 5), and NOS (*n* = 63). In summary, most ependymomas in our cohort did not contain ecDNA except for three patients, one of which matches the recently described molecular subgroup of aggressive *MYCN*-amplified spinal ependymoma.

*Choroid plexus tumors*

Choroid plexus tumors (CPT) are rare tumors derived from the choroid epithelium and comprise low-grade choroid plexus papillomas (CPP) and high-grade choroid plexus carcinomas (CPC). Of 26 CPCs and 32 CPPs in this cohort, we identified low-copy ecDNA in one CPC at chr20p11.2. There is currently little evidence to support oncogenic roles for any of the 7 intacct genes (*INSM1*, *KIZ*, *NKX2-2*, *NKX2-4*, *PAX1*, *RALGAPA2*, *XRN2*) amplified on this ecDNA (**Supplementary Table 5**); however, focal amplification of chr20p11.2 has previously been reported in breast cancer cell lines^3^ and in patient samples of other solid tumor types^4,5^.

*Pineal tumors*

Of 28 patients with pineal tumors, one pineoblastoma had ecDNA amplification of *MYC*. Methylation-based classification placed this tumor in the subgroup driven by *MYC*/*FOXR2* activation^6^. ecDNA was not detected in any of four low-grade rare pediatric pineal tumors. Given the limited number of samples included in this cohort, we expect that larger cohorts will be required to chart the genomic landscapes of ecDNA amplifications in choroid plexus and pineoblastoma tumors.

*Miscellaneous embryonal brain tumors*

Among 30 embryonal brain tumors not otherwise specified (NOS), we identified 4 ecDNA sequences mapping respectively to *C19MC* (including *TTYH1* fusion), chr12 (*CCND2*, *CDK4*, and *MDM2*), chr2 (*ALK*), and chr6 (*PLAGL1*). Embryonal tumors with *PLAGL* family amplification were recently proposed as a distinct molecular diagnosis^7^. In addition to the case classified here, we note 3 cases annotated as high-grade gliomas with ecDNA amplification of *PLAGL2* which may belong to this emerging entity.

*Meningioma*

We do not detect ecDNA in any of 61 pediatric meningiomas (0%). Focal amplification in meningiomas is rare^8^, although at least one case has previously been reported with ecDNA amplification of *MDM2*^9^.

**Supplementary Note 4**

*Other extracranial solid tumors with ecDNA.*

*Wilms tumor*

Wilms' tumors have diverse genetic drivers including *MYCN* amplification in some cases^10,11^. We identify ecDNA sequences in 4 of 63 tumors (6%), amplifying respectively *MYCN*; *MYCL*; chr20q11 including *BCL2L1*, *ID1*, *PLAGL2* and *SRC*; and segments of chr4p containing no known oncogenes.

*Adrenocortical carcinoma*

Adrenocortical carcinomas (ACC) are genetically heterogeneous and some harbor focal amplifications of driver genes, most commonly *TERT* and *CDK4*^12^. We identify 5 low-copy ecDNA amplifications in 4 of 23 tumors (17%), originating from cytogenetic bands chr2q23.3, chr14q32.33, chr8q21.11, chr11q13, and chr11p11.2. Only one of these sequences contained full-length known oncogenes (*INPPL1* and *NUMA1*). Given that we do not observe intra- or extrachromosomal amplification in this cohort of the above recurrently amplified drivers of ACC, we conclude that our sample is insufficient to represent the full spectrum of focal amplification in ACC.

*Germ cell tumors*

The 50 germ cell tumors (GCT) in this cohort comprised histological diagnoses including teratomas, yolk sac tumors, dysgerminomas and germ cell tumors originating from various tissues. Two tumors (4%) had ecDNA: one GCTNOS amplifying *MYCN* and a brain yolk sac tumor amplifying *MDM2*.

*Benign tumors*

The histological diagnoses classified as benign included adenomas, fibromas, cavernomas, neuromas, lipomas, lipoblastomas, osteomas, osteoblastomas, Langerhans Cell histiocytoses, and other rare tumors. One hepatic focal nodular hyperplasia was classified as ecDNA(+), containing a short (50kbp) intergenic low-copy amplification of chr2:57170434-57220435 (hg38).

*Peripheral nerve sheath tumors*

The 80 nerve sheath tumors in this dataset comprised 41 schwannomas, 3 perineuromas (PN), 1 adenomatoid odontogenic tumor, 23 neurofibromas plexiform (NFP), and 10 malignant peripheral neural sheath tumors (MPNST). Of these tumor types, only MPNST is malignant and may arise from lower-grade PN or NFP but not schwannomas^13^. Oncogenic amplifications are common in MPNST^14^ but whether these may be extrachromosomal remains poorly understood. We identify 2 MPNSTs with ecDNA and 2 with intrachromosomal but not extrachromosomal amplifications. In one case, the ecDNA sequence amplified segments of chr12 and chr13 of sum length 6.7Mbp including *CDK4* and *MDM2*. In the other case, we identify ecDNA in both longitudinal samples of the same MPNST tumor. The ecDNA sequences in these samples shared some minimal sequence similarity indicating a possible shared origin, but the amplified oncogenes differed: *TWIST1* in the first, and *MET* and *SMO* in the second sample (**Supplementary Table 10**). In summary, we find evidence of ecDNA only in malignant peripheral nerve sheath tumors, and not in benign tumors of the same tissues.

*Miscellaneous rare pediatric sarcomas*

Other sarcomas with ecDNA comprised a spindle cell sarcoma with two ecDNA sequences, one amplifying *KIT* and *PDGFRA* oncogenes and the other amplifying no known oncogenes but genes with plausible roles in cancer including *APIP*, *CAT*, and the immune regulator *CD44*; a sarcoma not otherwise specified with high-copy ecDNA amplification of *MYC*, and 4 sarcomas with ecDNA containing no known oncogenes and of unknown prognostic relevance.

*Hepatoblastoma*

Hepatoblastomas (HBL) are rare childhood tumors of the developing liver. Recurrent focal amplifications of chr2q24.3 have previously been described^15^ but the exact oncogenic sequence is not known, nor whether this amplification may be extrachromosomal. We found ecDNA in 2 of 14 HBL tumors (14%), both amplifying chr2q24.3. The minimal region amplified on both ecDNA sequences spanned 5Mbp including 19 protein-coding genes, none with known oncogenic roles. No other tumor type amplified segments of chr2q24.3 containing complete gene loci, suggesting that a possible oncogenic role for this locus may be specific to hepatoblastoma. Further investigation is warranted in a tumor-specific cohort to elucidate the exact oncogenic sequence and its role in tumorigenesis.

*Melanoma*

Pediatric melanomas are much rarer than their adult counterparts. Copy number variation in pediatric cases is well-described, but the extent of focal amplification, especially extrachromosomal, is not^16^. We identify high-copy, chromothriptic ecDNA in one case out of 13 (8%). The computationally reconstructed amplification spanned segments of chromosomes 3, 4, 7, 12, and 19 and included known oncogenes *ACTN4*, *AKT2*, *AXL*, *RAC1*, *SERTAD1*, and *SERTAD3*.

**Supplementary Note 5**

*Analysis of pathogenic germline variation with respect to ecDNA in pediatric central nervous system tumors.*

Pediatric tumors have higher rates of pathogenic germline driver variation than adult cancers^17^, highlighting an opportunity to investigate whether germline cancer predisposition syndromes may be associated with later development of tumors with ecDNA. To explore possible association between pathogenic germline variation and focal amplification including ecDNA, we reanalyzed high-quality annotations of oncogenicity available for 786 genomes in the OpenPBTA subcohort^18^, using a similar approach to that applied to analyze pathogenic simple somatic variants. Excluding tumor types without ecDNA yielded an analysis set of 441 pediatric CNS tumor cases. Known oncogenic germline simple variants were aggregated at the gene level, yielding 18 testable genes. For each, association was first evaluated with respect to focal amplification of any kind, and then, within the amplified subset, with ecDNA specifically (*χ*^2^ test).

Focally amplified tumors were enriched for germline pathogenic *TP53* variation (Li-Fraumeni Syndrome, LFS; *χ*^2^ = 28, *p* = 1e-7), but no specific association between germline *TP53* alteration and ecDNA was detected. This is consistent with known links between genomic instability, *TP53* inactivation, and ecDNA^19,20^; and with evidence linking LFS specifically to genomic instability^21,22^, although the relative propensity for LFS tumors to develop focal amplification or ecDNA remains to be determined. We note, however, that despite their relative enrichment in pediatric cancers, germline drivers still account for only a small minority of tumors in this subcohort (12%)^18^, limiting statistical power for discovery. We anticipate that future investigations of ecDNA across larger datasets will enable more definitive assesment of ecDNA in cases with germline cancer predisposition syndromes.

13.	Gilchrist, J. M. & Donahue, J. E. Peripheral nerve tumors. in *UpToDate* (ed. Connor, R. F.) (Wolters Kluwer, 2024).
